# Consensus recommendations for measuring the impact of contraception on the menstrual cycle in contraceptive clinical trials

**DOI:** 10.1101/2024.04.04.24305350

**Authors:** Amelia C. L. Mackenzie, Stephanie Chung, Emily Hoppes, Nora Miller, Anne E. Burke, Sharon L. Achilles, C. Leigh Allen, Luis Bahamondes, Diana L. Blithe, Vivian Brache, Rebecca L. Callahan, Alice F. Cartwright, Kathryn B. H. Clancy, Enrico Colli, Amanda Cordova-Gomez, Elizabeth E. Costenbader, Mitchell D. Creinin, Hilary O. D. Critchley, Gustavo F. Doncel, Laneta J. Dorflinger, Alison Edelman, Thomas Faustmann, Christoph Gerlinger, Lisa B. Haddad, Julie Hennegan, Cássia Raquel Teatin Juliato, Simon P. S. Kibira, Diana Mansour, Andres Martinez, Kristen A. Matteson, Jacqueline A. Maybin, Alexandria K. Mickler, Kavita Nanda, Chukwuemeka E. Nwachukwu, Funmilola M. OlaOlorun, Kevin J. Peine, Chelsea B. Polis, Carolina Sales Vieira, Regine Sitruk-Ware, Jennifer A. Smit, Marsden Solomon, Lisa M. Soule, Douglas Taylor, Elizabeth E. Tolley, Olivia Vandeputte

## Abstract

**Objective:** We sought to develop consensus recommendations for improved measurement of contraceptive-induced menstrual changes (CIMCs) in contraceptive clinical trials to build upon previous standardization efforts.

**Study Design:** We undertook a virtual consensus-building process using a modified Delphi approach, incorporating aspects of Nominal Group Technique and the Jandhyala method. The process consisted of four half-day meetings, developing recommendations within five topical working groups, a series of consensus questionnaires to assess recommendation agreement on a 4-point Likert-like scale, and opportunities for input and feedback throughout the process. Recommendations required at least 75% consensus to be included.

**Results:** Our interdisciplinary group—from 26 organizations and 13 countries in five global regions across academia, nonprofit research organizations, the pharmaceutical industry, and funding agencies— developed 44 consensus recommendations. These included recommendations on standardization, patient-reported outcome measure development aligned with current regulatory guidance, trial design, protocol development, participant recruitment, data collection, data analysis, and areas for exploratory research. Seven recommendations were deemed a priority by over 85% of our group.

**Conclusion:** Using a formal consensus-building process, we reached high levels of agreement around recommendations for more standardized and patient-centered measurement of CIMCs in contraceptive clinical trials, now and in the future.

**Implications statement:** Consensus recommendations on measuring bleeding changes and related outcomes in contraceptive clinical trials can improve reporting of standardized, patient-centered outcomes in future product labeling. These improvements can enable providers to offer more relevant counseling on contraceptives and permit greater comparability and data synthesis across trials to inform clinical guidance.

## 1. INTRODUCTION

The use of contraception can cause changes in uterine bleeding patterns, uterocervical fluid, and uterine cramping, and it can impact how users experience menstrual and gynecologic disorders and symptoms. Collectively, these changes can be referred to as contraceptive-induced menstrual changes (CIMCs), although there are differing perspectives on this terminology (Figure 1). Experiencing CIMCs can both negatively and positively impact use dynamics, health, and wellbeing for contraceptive users, as well as the individual and sociocultural acceptability of contraception [1]. When users find CIMCs undesirable, negative consequences can include contraceptive dissatisfaction, reduced quality of life, increased burden around managing menstrual health, reduced sexual wellness, and potential unintended pregnancy if users discontinue contraception while still wanting to prevent pregnancy [1–4]. On the other hand, experiences of CIMCs users consider desirable can result in benefits, such as management of for menstrual and gynecologic disorders and symptoms, method satisfaction, improved sexual wellness, and reduced burden or costs of menstrual materials [1–6]. Which CIMCs users determine to be desirable or undesirable for them can vary widely and is influenced by individual preferences and norms and community-level norms—especially around menstruation, menstrual health, and sexual and reproductive health—as well as perspectives of partners and wider social and contextual factors [1–3,7,8]. However, broadly, users often prefer reduced bleeding and cramping, and dislike increased bleeding and unpredictability [1,2].

**Figure 1.**
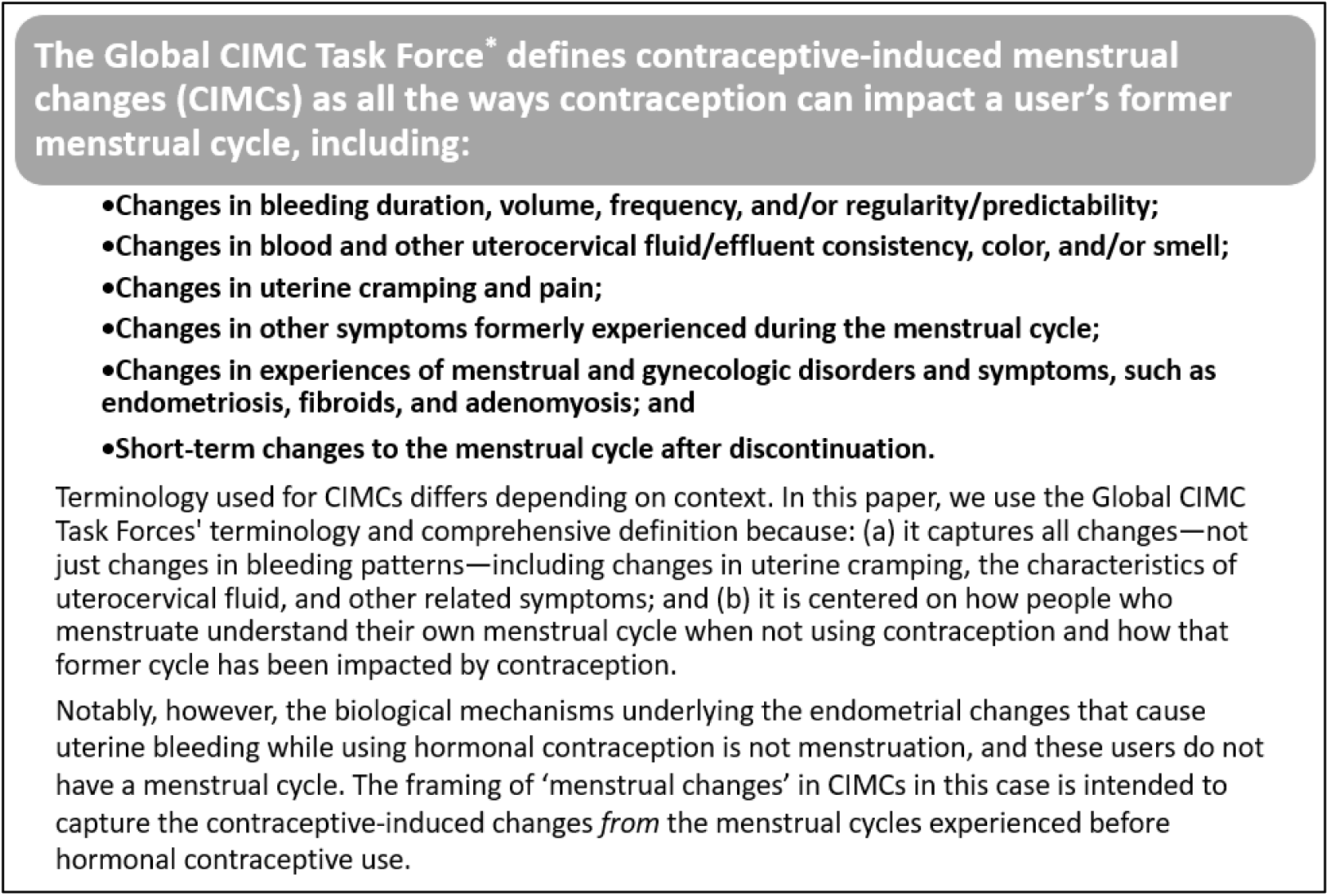
Terminology and definition of contraceptive-induced menstrual changes (CIMCs) *Citation: Hoppes E, Nwachukwu C, Hennegan J, Blithe DL, Cordova-Gomez A, Critchley H, Doncel GF, Dorflinger LJ, Haddad LB, Mackenzie ACL, Maybin JA, Moley K, Nanda K, Vieira CS, Vwalika B, Kibira SPS, Mickler A, OlaOlorun FM, Polis CB, Sommer M, Williams KM, Lathrop E, Mahajan T, Rademacher KH, Solomon M, Wilson K, Wilson LC, Rountree L. Global research and learning agenda for building evidence on contraceptive-induced menstrual changes for research, product development, policies, and programs. Gates Open Res. 2022 Apr 19:6:49. doi: 10.12688/gatesopenres.13609.1.

Considerations for CIMCs have played a notable role in contemporary contraceptive research and development (R&D). Beginning in the 1950s with the preclinical and clinical research for the first oral contraceptive, a number of development decisions were made, at least in part, due to CIMCs. For example, mestranol and a regimen of hormone-free days were both added because researchers decided monthly withdrawal bleeding was the most acceptable to contraceptive users, as well as possibly other contemporary influential stakeholders [9–11]. These decisions, made decades ago, still impact the current contraceptive method mix and contraceptive R&D today. Throughout this contemporary development of new contraceptives, there have been several endeavors to standardize how CIMCs are measured and reported, which we summarize in Figure 2. Creinin and colleagues have recently reviewed this history and the measurement outcomes from that work in detail [12]; therefore, we focus on the contributors and processes of these previous efforts to highlight differences from the approach reported in this paper.

**Figure 2.**
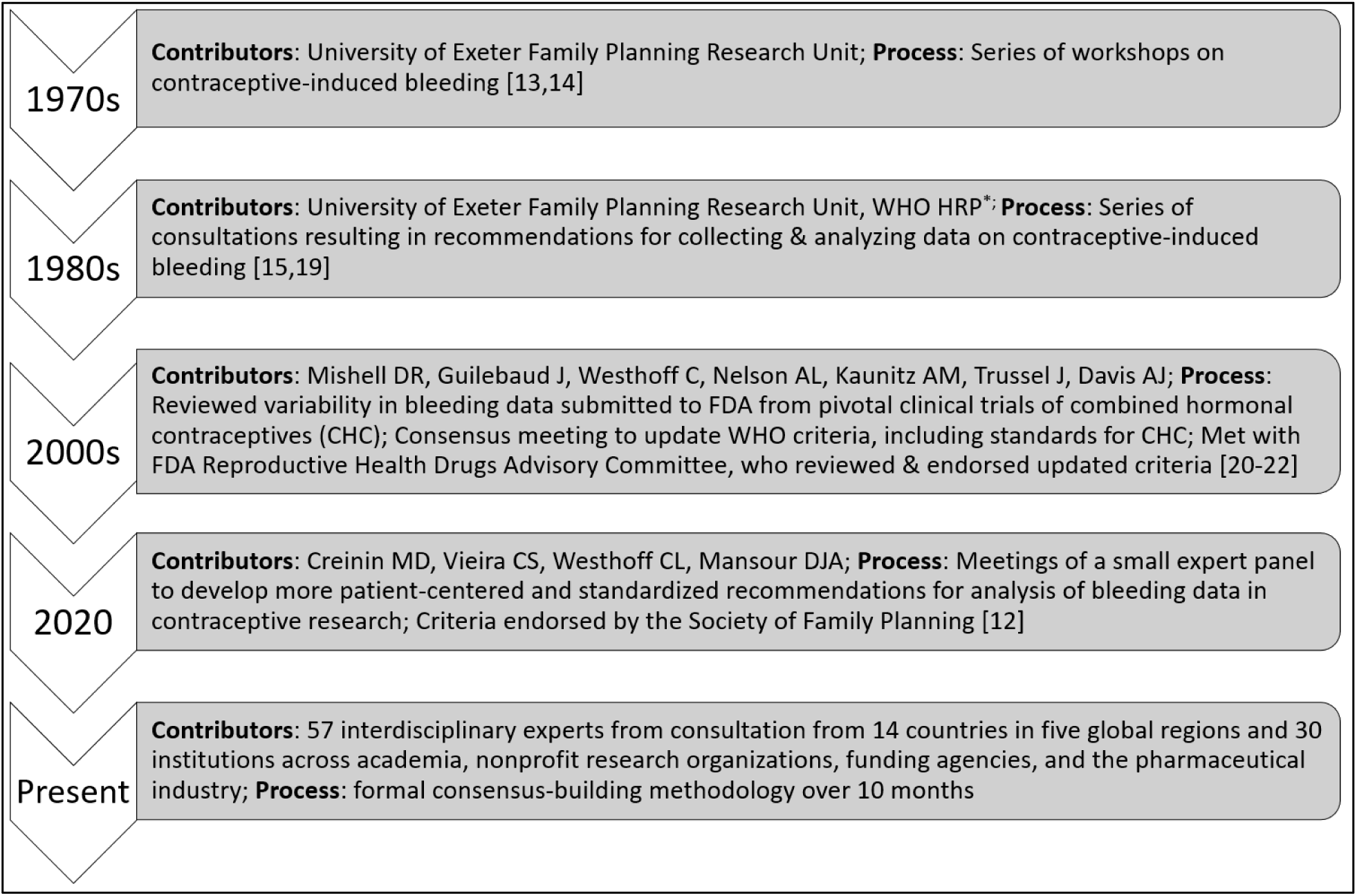
Overview of efforts to standardize the measurement and analysis of CIMC data in contraceptive clinical trials. *\* Special Programme of Research, Development and Research Training in Human Reproduction* [23]

Current practices for measuring CIMCs in trials continue to have limitations, including burdensome data collection, a lack of uniform definitions provided to trial participants to understand key terms, and the use of patient-reported outcome (PRO) data without evidence of validation in line with current regulatory guidance on patient-focused drug development [24–27]. These limitations are particularly relevant for standard measurement of CIMCs in trials globally because—for topics like menstruation, with high levels of stigma—there are generally low levels of shared understanding and common references within and between communities and contexts [8,28,29]. In addition, how CIMC outcomes reported by trials for providers to use in contraceptive counseling of new methods are often not meaningful to contraceptive users to facilitate their informed method decision-making. Building upon previous standardization efforts—and in collaboration with many of the experts who contributed to this earlier work—we developed consensus recommendations to further improve how CIMCs are measured in contraceptive clinical trials now and in the future. Our consensus-building approach to address current measurement limitations focused on: (a) incorporating a variety of interdisciplinary and global perspectives, (b) considering current regulatory guidance around PROs, and (c) including all types of CIMCs, not just changes in bleeding patterns, to align with the Global CIMC Task Force’s comprehensive definition.

## 2. METHODS

Members of a scientific and technical team at FHI 360 engaged a group of interdisciplinary experts about participating in a consensus-building process to improve how CIMCs are measured in contraceptive clinical trials. Invitees had a range of expertise in topics (i.e., contraceptive clinical trials, family planning, the menstrual cycle, menstrual health, measurement and analysis approaches, and regulatory considerations) and disciplines (i.e., clinical research, social-behavioral science, biomedical research, basic science, and clinical practice). Experts were recruited within multiple sectors from members of the existing CIMC Global Task Force [1], contributors to previous standardization efforts, contributors to the literature in relevant fields, and suggestions from other experts. Among these experts, we convened a 13-member external planning committee from 12 organizations and 6 countries who volunteered to contribute to defining objectives for the expert consultation and refining the proposed consensus-building process.

Our objectives were: (a) to review and discuss limitations and strengths of current approaches for measuring and analyzing data on CIMCs in contraceptive clinical trials; (b) to come to a consensus on recommendations for improvements in CIMC measurement and analysis in contraceptive clinical trials that meet the needs of trial participants, researchers, sponsors, regulators, and future users; and (c) to identify important, related topics outside the scope of this expert consultation that warranted similar consideration. The second objective also included determining what research and evidence are needed to empirically evaluate the recommended improvements. Our consensus-building methodology was an amalgamation of the RAND/UCLA method, the Delphi method, the Modified Rand-Delphi method, the Nominal Group Technique, and the Jandhyala method because no single approach met all our needs [30–35]. We present extensive details on the methodology in the Supplementary Material, but briefly, the fully virtual consensus-building process, depicted in Figure 3, included four half-day meetings for all experts to join, recommendation development within five smaller topical working groups into which experts self-selected, and a series of consensus questionnaires around the scope and details of the recommendations.

**Figure 3.**
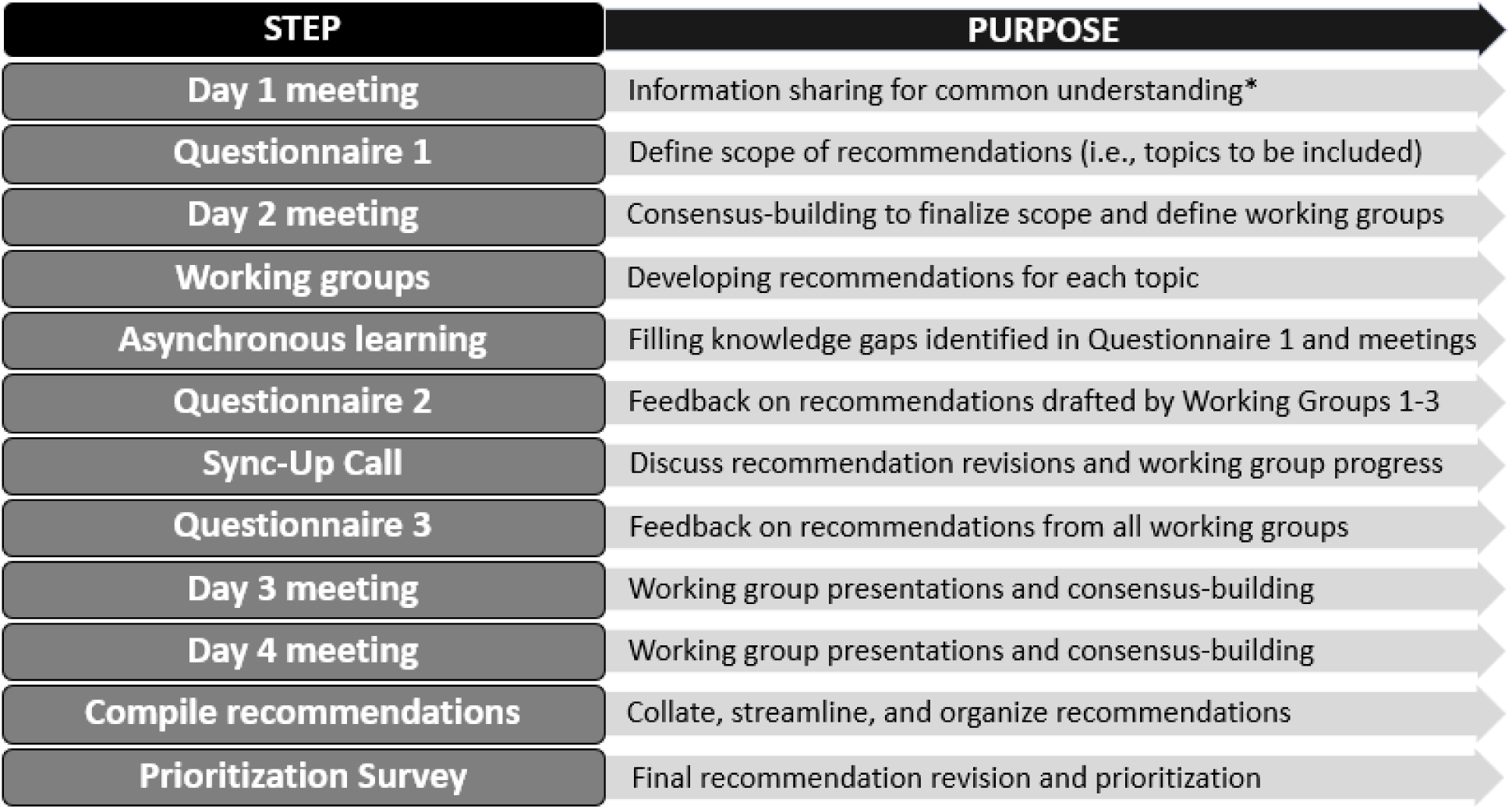
Consensus-building methodology for developing recommendations on CIMC measurement in contraceptive clinical trials. *\*To establish a shared understanding on key information within our interdisciplinary group, including on contraceptive clinical trials, patient-reported outcome measures, sociocultural contexts and CIMCs, and inclusive language around gender and sexuality*

Each consensus questionnaire asked experts about their agreement/disagreement with recommendations on a four-point Likert-like scale (i.e., strongly agree, somewhat agree, somewhat disagree, strongly disagree) and included at least one open-ended question for additional input. One author (SC) completed qualitative analysis of open-ended questions. During the first two days of meetings, we decided topics would need 85% agreement (i.e., ‘strongly agree’ or ‘somewhat agree’) among experts to be considered within scope of this work, and specific recommendations within those topics would need 75% agreement among members of working groups to be included in our final list. The Office of International Research Ethics at FHI 360 determined the expert consultation protocol was research that was exempt from Institutional Review Board review.

## 3. RESULTS

### 3.1. Expert characteristics

A total of 57 experts were involved in the expert consultation from 30 organizations—across academia, nonprofit research organizations, funding agencies, and the pharmaceutical industry—and 14 countries in five global regions (i.e., Northern America, Europe, Africa, Latin America & the Caribbean, and Oceania, listed in order of frequency). Of those experts, 44 self-selected into working groups. Working group members represented over 625 combined years of experience—including nearly 70% with more than 15 years—in contraceptive clinical trials and other R&D, menstrual cycle and menstrual health research, social-behavioral research, contraceptive prescribing, and/or other relevant fields (Table 1). Most had doctoral training in either obstetrics and gynecology and/or research and worked either in academia or at nonprofit research organizations. Most experts self-identified as women, and about half were under the age of 50 and half were 50 or more years old. Almost all experts (88%) reported having either experienced CIMCs themselves or observed someone personally close to them experiencing CIMCs.

**Table 1:**
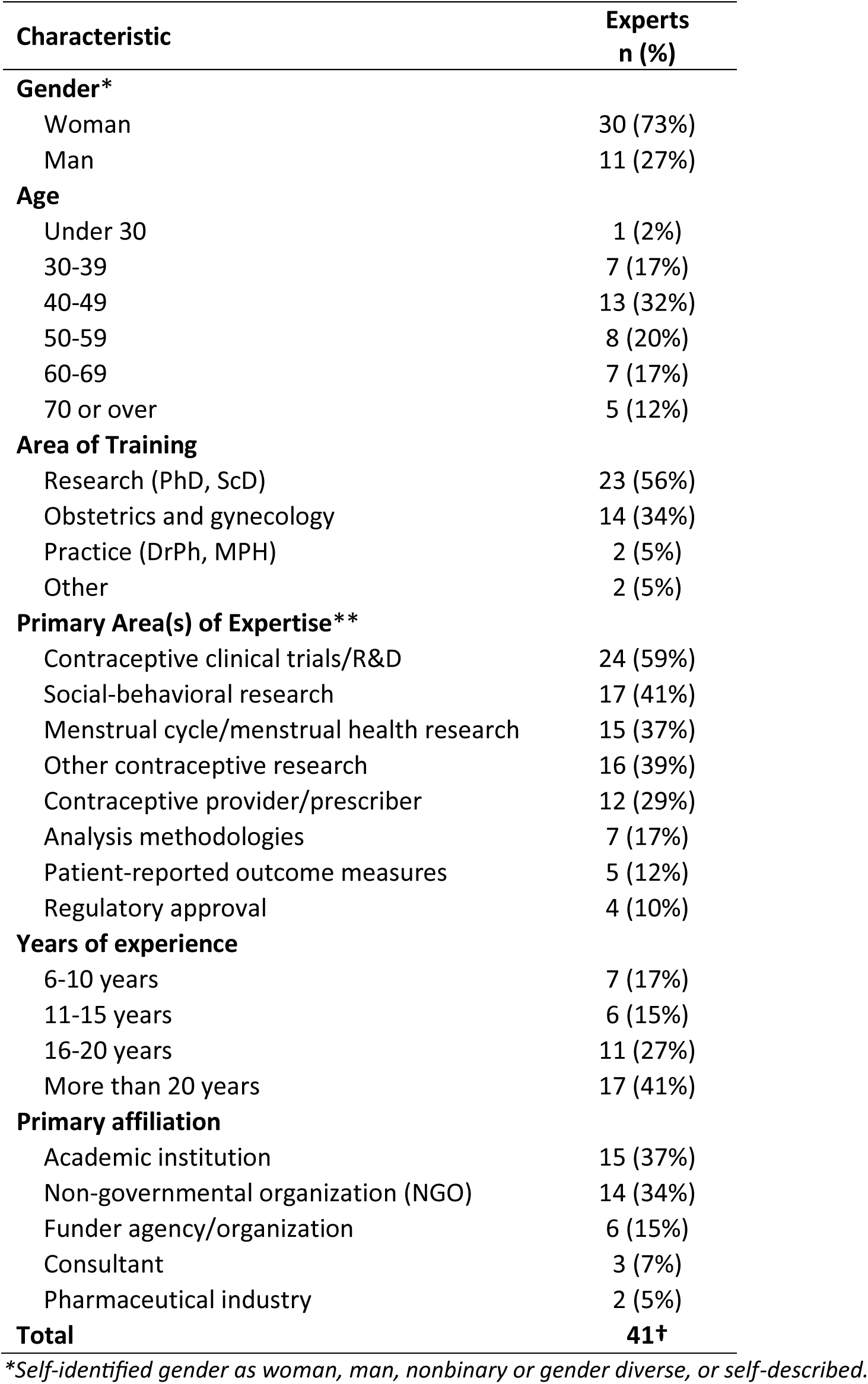
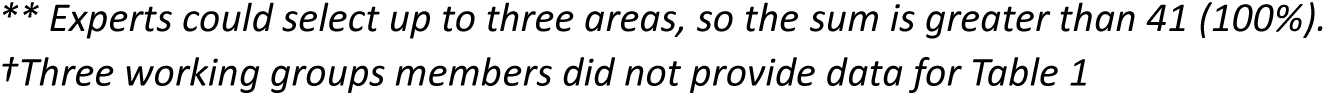
Characteristics of experts in topical working groups developing recommendations on CIMC measurement in contraceptive clinical trials.

| **Characteristic** | **Experts**<br>**n (%)** |
| --- | --- |
| **Gender\*** |  |
| Woman | 30 (73%) |
| Man | 11 (27%) |
| **Age** |  |
| Under 30 | 1 (2%) |
| 30-39 | 7 (17%) |
| 40-49 | 13 (32%) |
| 50-59 | 8 (20%) |
| 60-69 | 7 (17%) |
| 70 or over | 5 (12%) |
| **Area of Training** |  |
| Research (PhD, ScD) | 23 (56%) |
| Obstetrics and gynecology | 14 (34%) |
| Practice (DrPh, MPH) | 2 (5%) |
| Other | 2 (5%) |
| **Primary Area(s) of Expertise\*\*** |  |
| Contraceptive clinical trials/R&D | 24 (59%) |
| Social-behavioral research | 17 (41%) |
| Menstrual cycle/menstrual health research | 15 (37%) |
| Other contraceptive research | 16 (39%) |
| Contraceptive provider/prescriber | 12 (29%) |
| Analysis methodologies | 7 (17%) |
| Patient-reported outcome measures | 5 (12%) |
| Regulatory approval | 4 (10%) |
| **Years of experience** |  |
| 6-10 years | 7 (17%) |
| 11-15 years | 6 (15%) |
| 16-20 years | 11 (27%) |
| More than 20 years | 17 (41%) |
| **Primary affiliation** |  |
| Academic institution | 15 (37%) |
| Non-governmental organization (NGO) | 14 (34%) |
| Funder agency/organization | 6 (15%) |
| Consultant | 3 (7%) |
| Pharmaceutical industry | 2 (5%) |
| **Total** | **41†** |

\*Self-identified gender as woman, man, nonbinary or gender diverse, or self-described.
**\*\* Experts could select up to three areas, so the sum is greater than 41 (100%).**
**†Three working groups members did not provide data for Table 1**

### 3.2. Defining the scope for recommendation development

All 11 topics proposed in Questionnaire 1 reached consensus for inclusion (Table 2). Experts suggested another five topics that did not reach consensus during voting during the Day 2 meeting, but a number of elements of those topics were incorporated into other consensus topics. Topics determined to be out of scope are presented in Supplementary Table S1, aligning with the third objective of the expert consultation to identify important, related topics that warrant similar consideration outside of the present process. We converged the 11 topics into 3 cross-cutting themes and 5 topical working groups (Table 2). Working group scopes were: (1) **Eligibility**, enrollment requirements, and confounders; (2) Type, frequency, and format/mode of CIMC **data** collected; (3) Measurement of **acceptability**, impact on daily life, culture, and context; (4) Research agenda for developing CIMC **instruments**; (5) **Analysis** methodology, measures, and analyses standardization.

**Table 2.** Scope of topics for recommendations on CIMC measurement in contraceptive clinical trials, as determined via Questionnaire 1 and Day 2 meeting, and the corresponding topical working groups.

| Topics from Questionnaire 1 | Percent agreement | Related working group |
| --- | --- | --- |
| Eligibility and enrollment requirements | 92% | Working Group 1. Eligibility |
| Data collection on confounders and other variables | 95% |  |
| Frequency of CIMC data collection | 100% | Working Group 2. Data |
| CIMC instrument format and mode | 95% |  |
| Type of CIMC data collected during clinical trials | 100% |  |
| Measurement of acceptability and impact on daily life | 100% | Working Group 3. Acceptability |
| Culture and context | 87% |  |
| Developing new CIMC instruments | 85% | Working Group 4. Instruments |
| Measures and analyses standardization | 97% | Working Group 5. Analysis |
| Analysis methodology | 85% |  |
| Regulatory considerations | 85% | Cross-Cutting Themes |
| Culture and context | 87% |  |
| Improved counseling information *† | 56% |  |
| Guiding frameworks (reproductive justice, life course, equity) * | 47% | Did not reach consensus during Day 2 voting. |
| Trial sample characterization (life stage, fertility intentions, goals/expectations, desires/needs) * | 50% |  |
| Non-contraceptive use (label information, menstrual choice, impact on wider health) * | 47% |  |
| Menstrual changes data in other trials beyond contraception * | 47% |  |

**\* Suggested by experts in Questionnaire 1, with percent agreement from Day 2 voting**
**† Revised into “Incorporating User Perspectives and Understandings of CIMCs” and agreed upon as cross-cutting theme**

### 3.3. Final Consensus Recommendations

We organized the final 44 consensus recommendations into six sections on: (a) *Standardization* in recommendations 1.1 to 1.4; (b) *Instruments for measuring CIMCs* in 2.1 to 2.3; (c) *Trial design, protocol development, and participant recruitment* in 3.1 to 3.8; (d) *Data collection* in 4.1 to 4.13; (e) *Data analysis* in 5.1 to 5.14; and (f) *Areas for exploratory research* in 6.1 and 6.2 (Table 3). We reached consensus on all recommendations, with nearly half (n=21) achieving greater than 90% consensus and six reaching 100% consensus (see Supplementary Table S2 for full consensus results). One aspect of one recommendation did not reach the consensus threshold of 75% by the end of our process (i.e., part of 5.3 on variables for analysis, details in Table S2), so it was not included. During the final stage of the consensus-building process (Figure 3), we identified priority recommendations based on the urgency and potential impact of their implementation (see Supplementary Table S2 for additional details on consensus and prioritization outcomes). This included seven recommendations that over 85% of experts deemed to be a priority and an additional 12 that at least 75% deemed a priority (highlighted by “*” or “†”, respectively in Table 3).

**Table 3.**
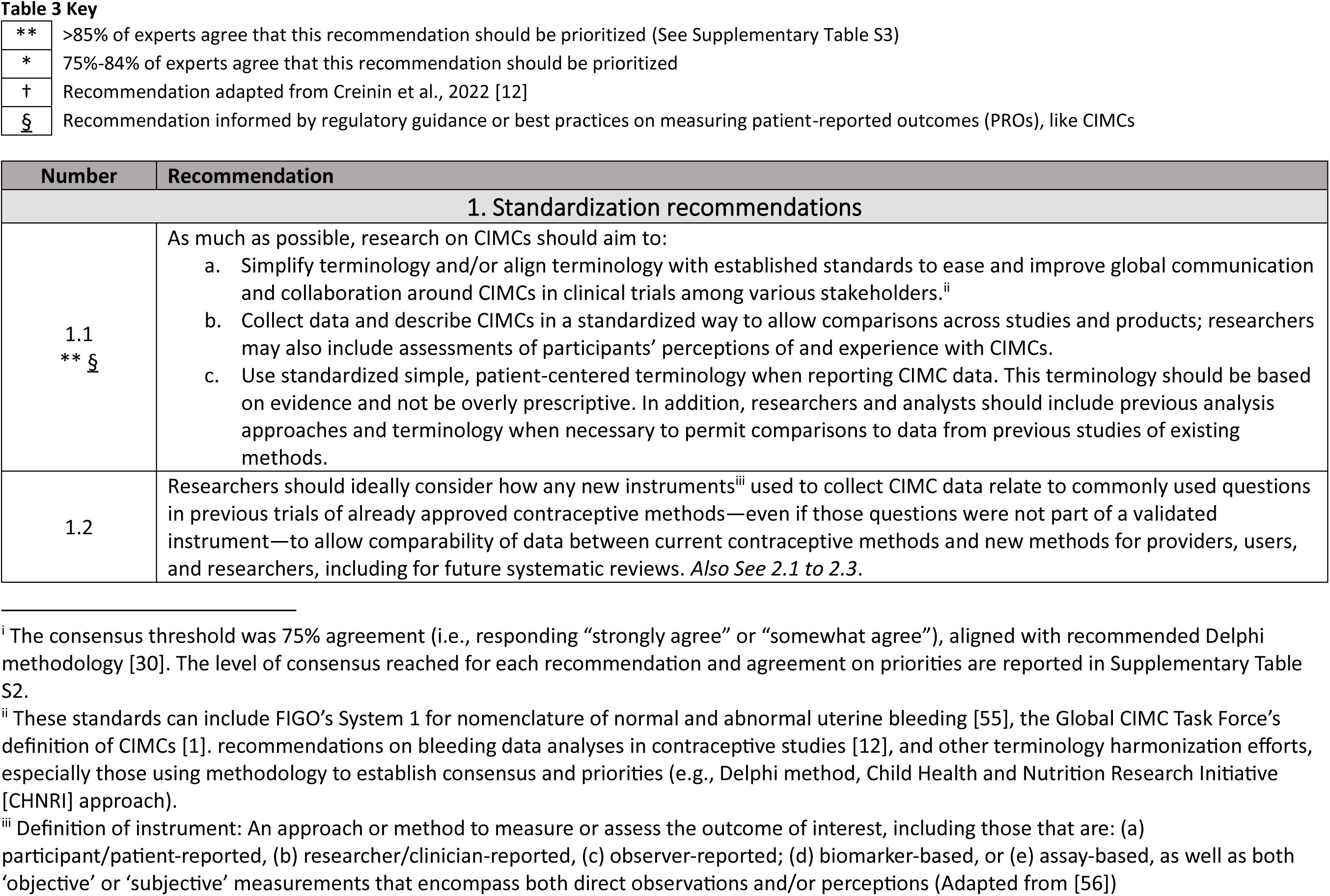
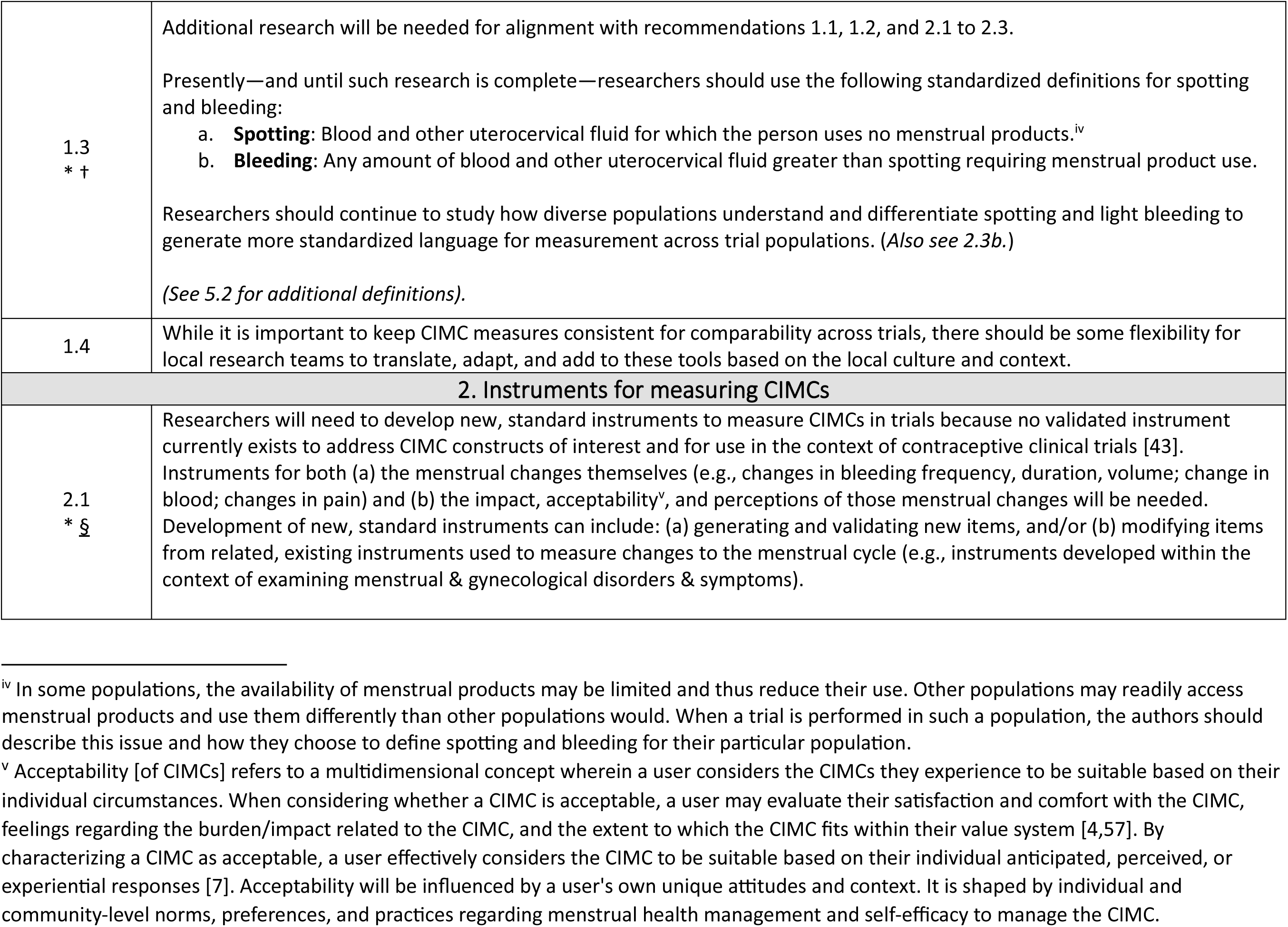
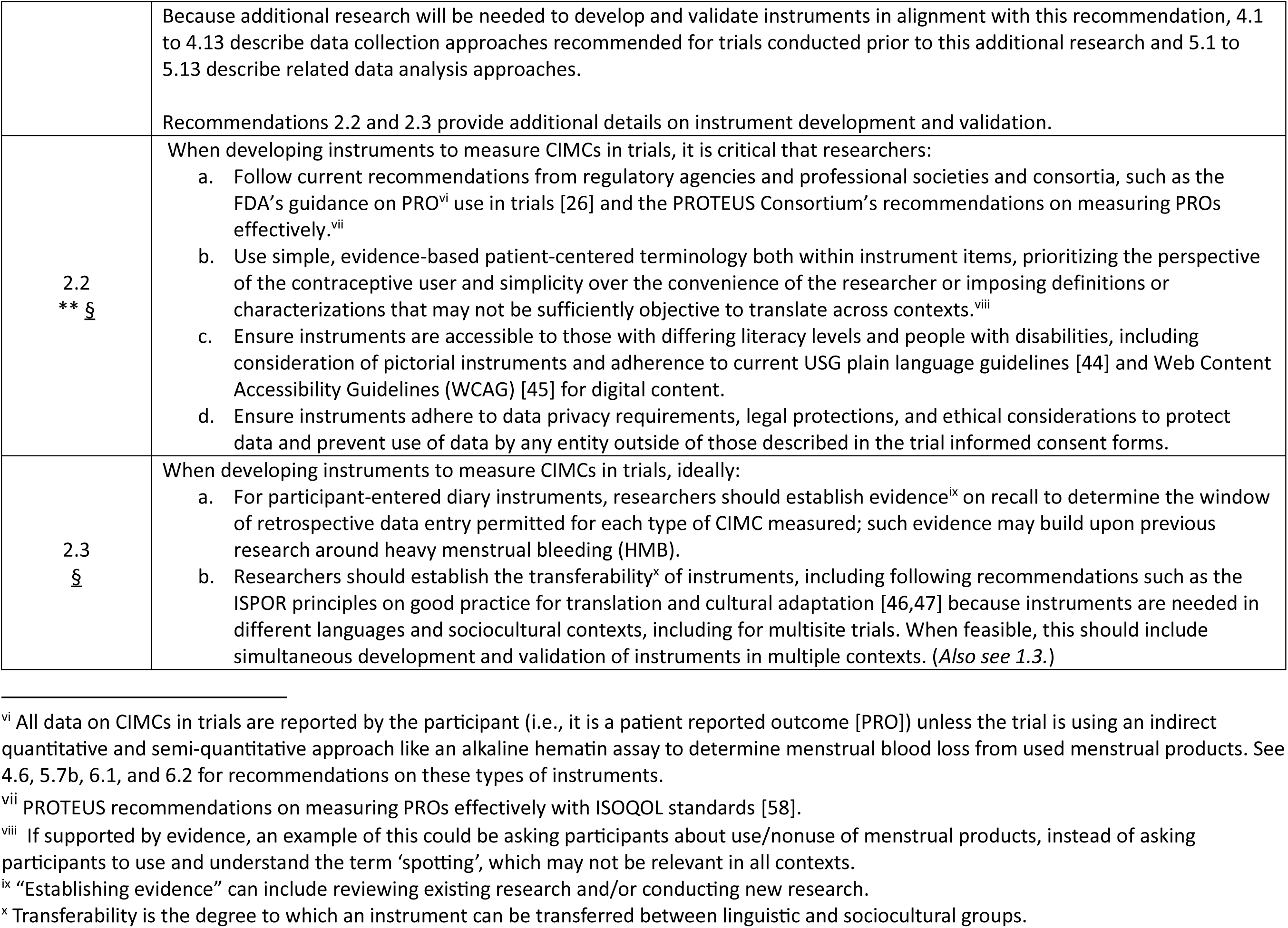
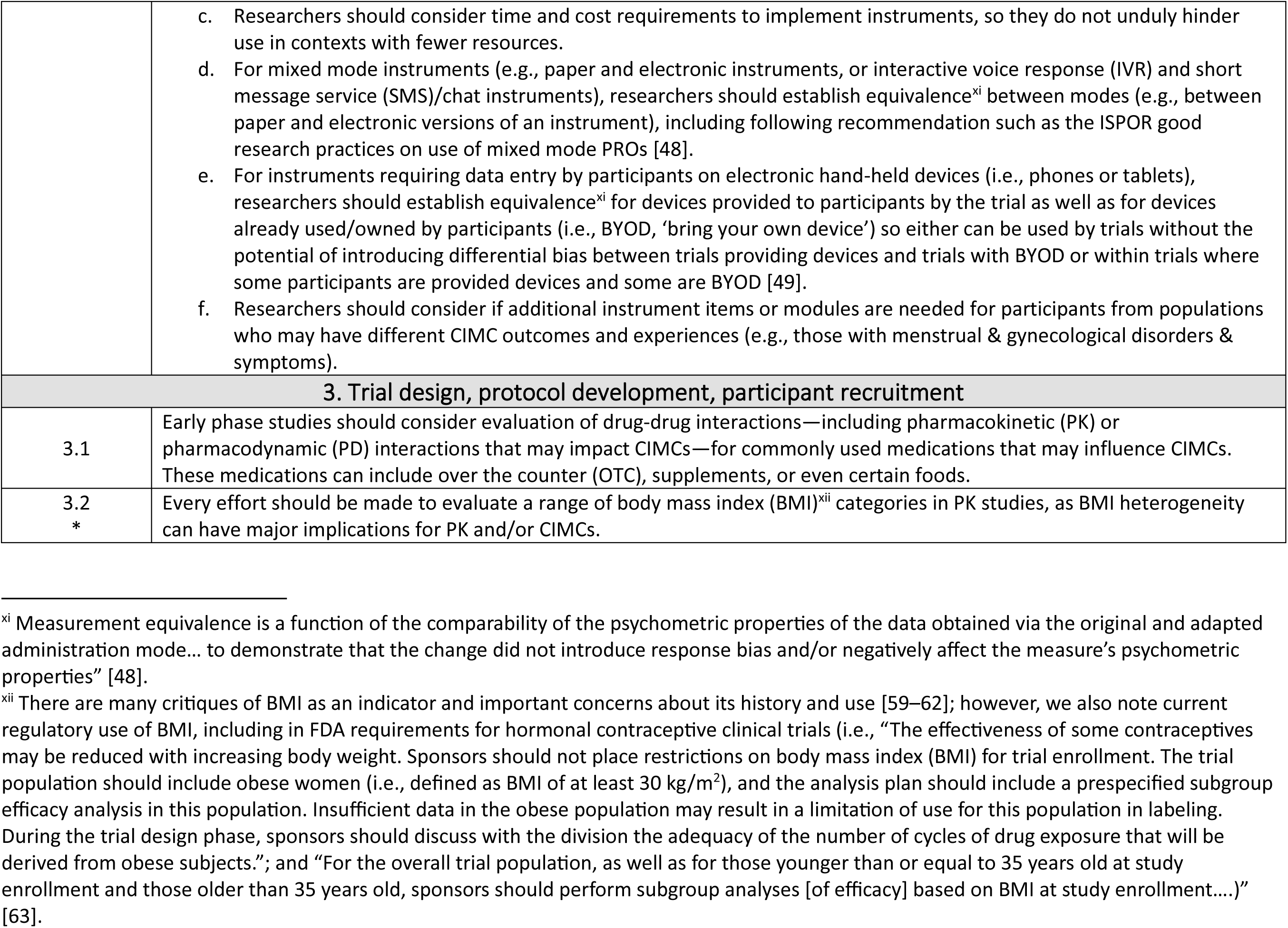
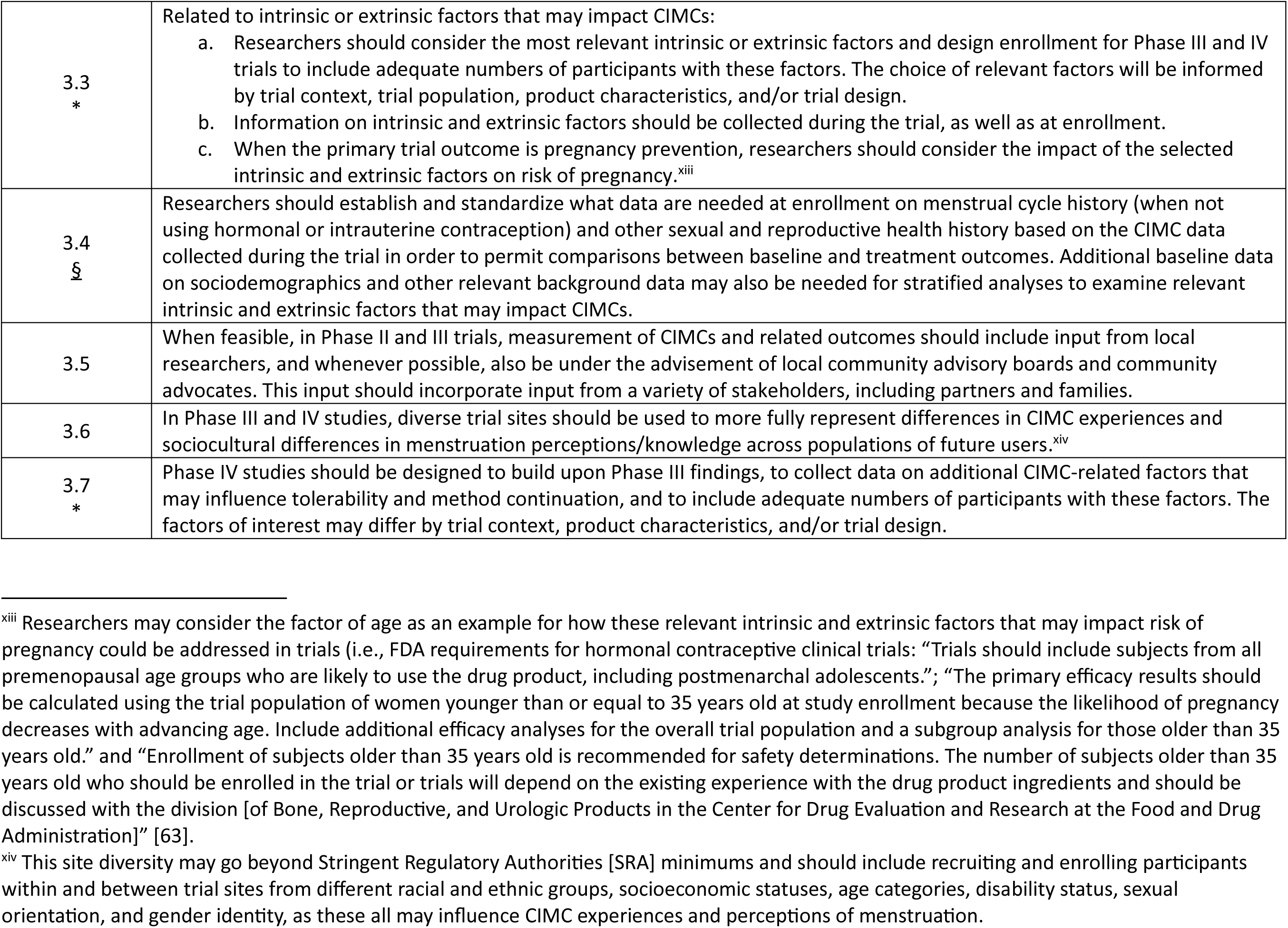
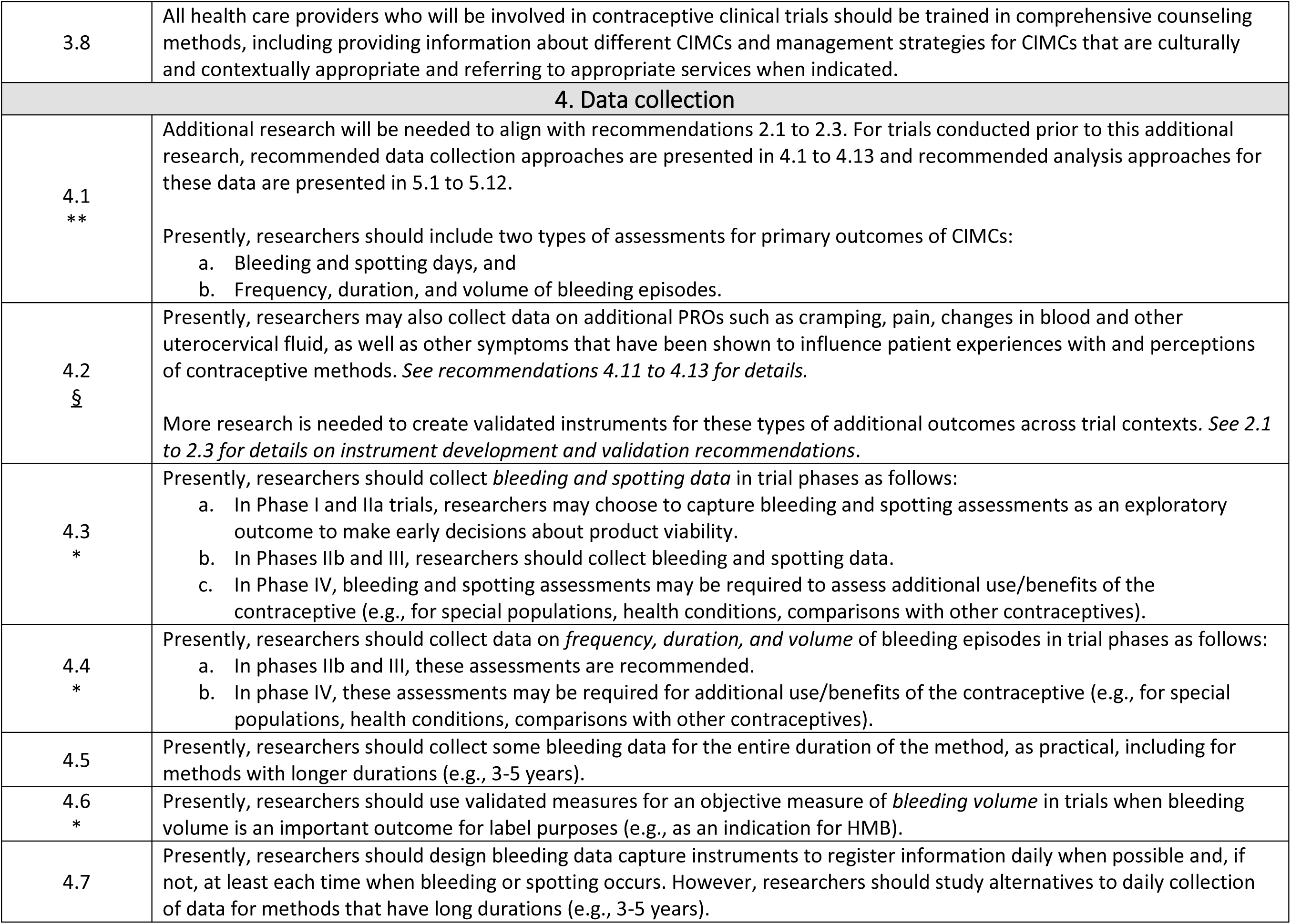
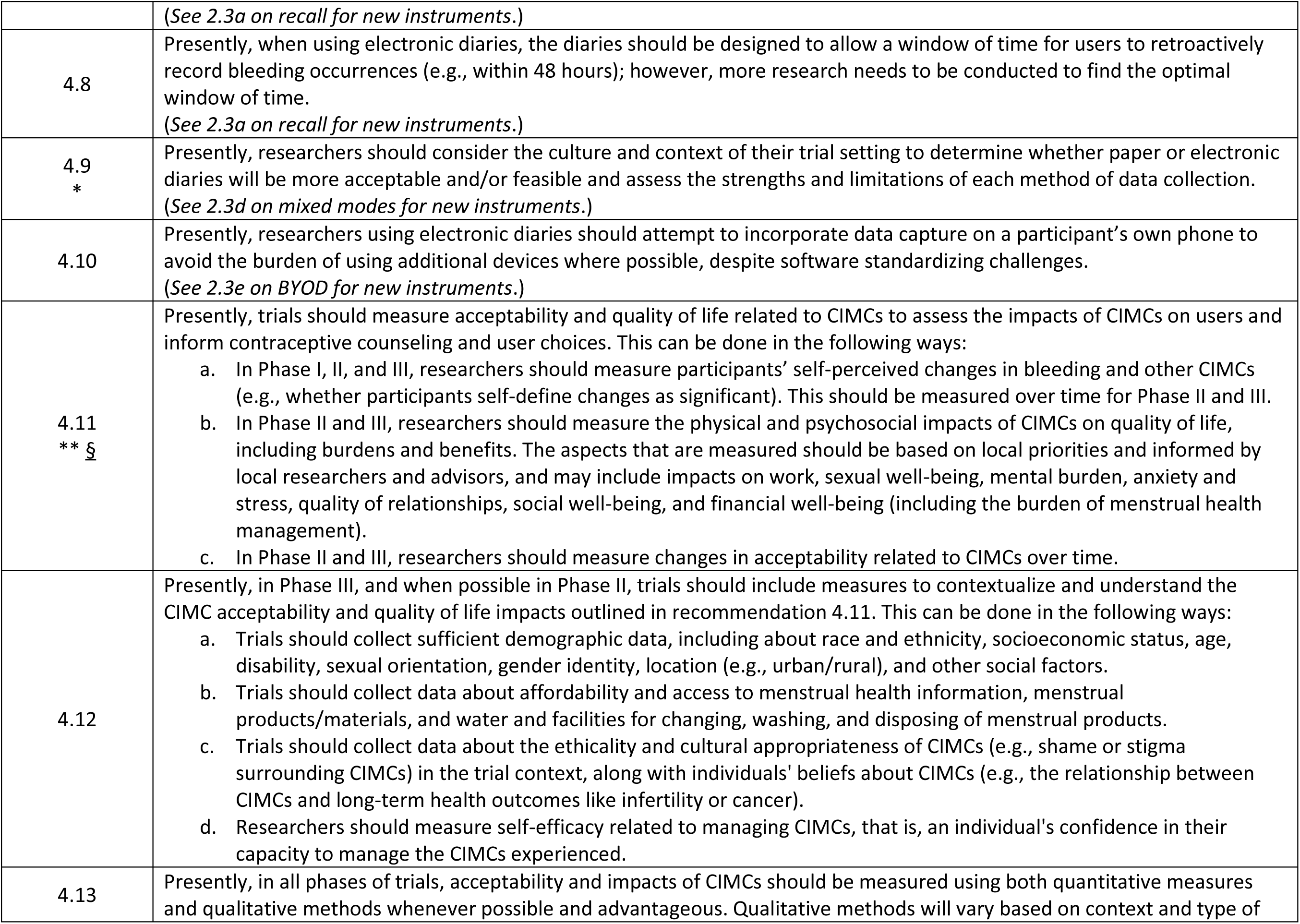
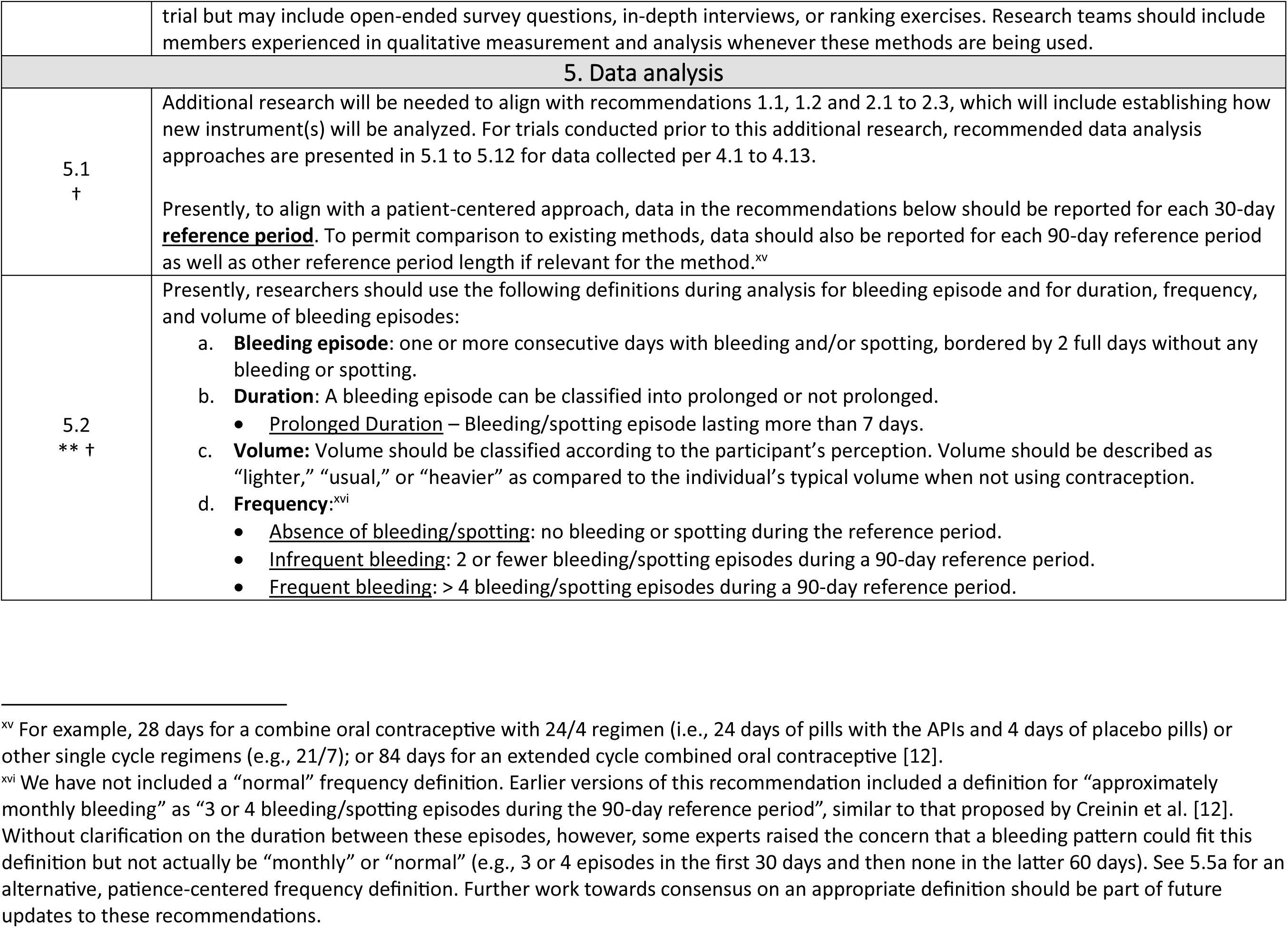
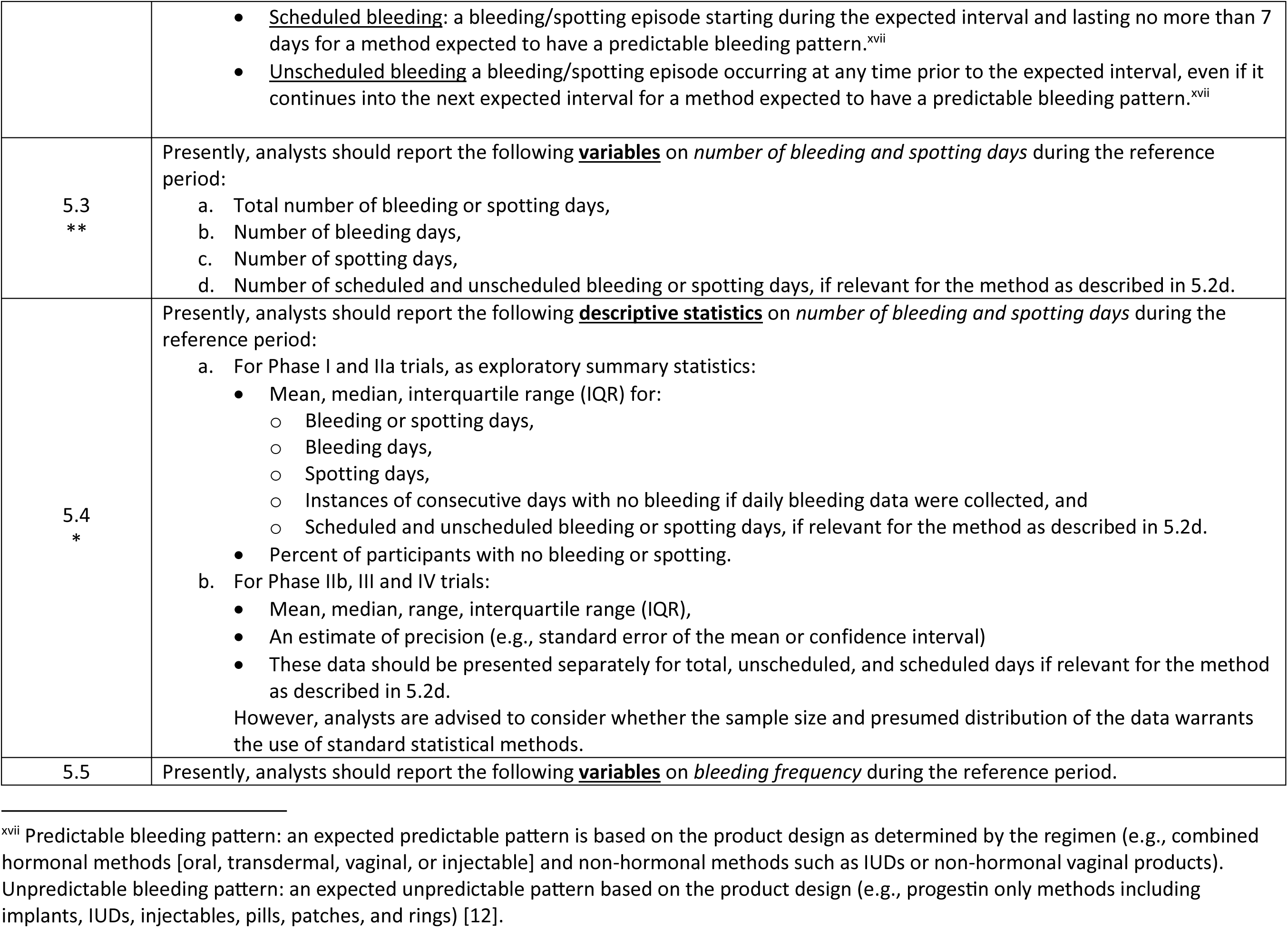
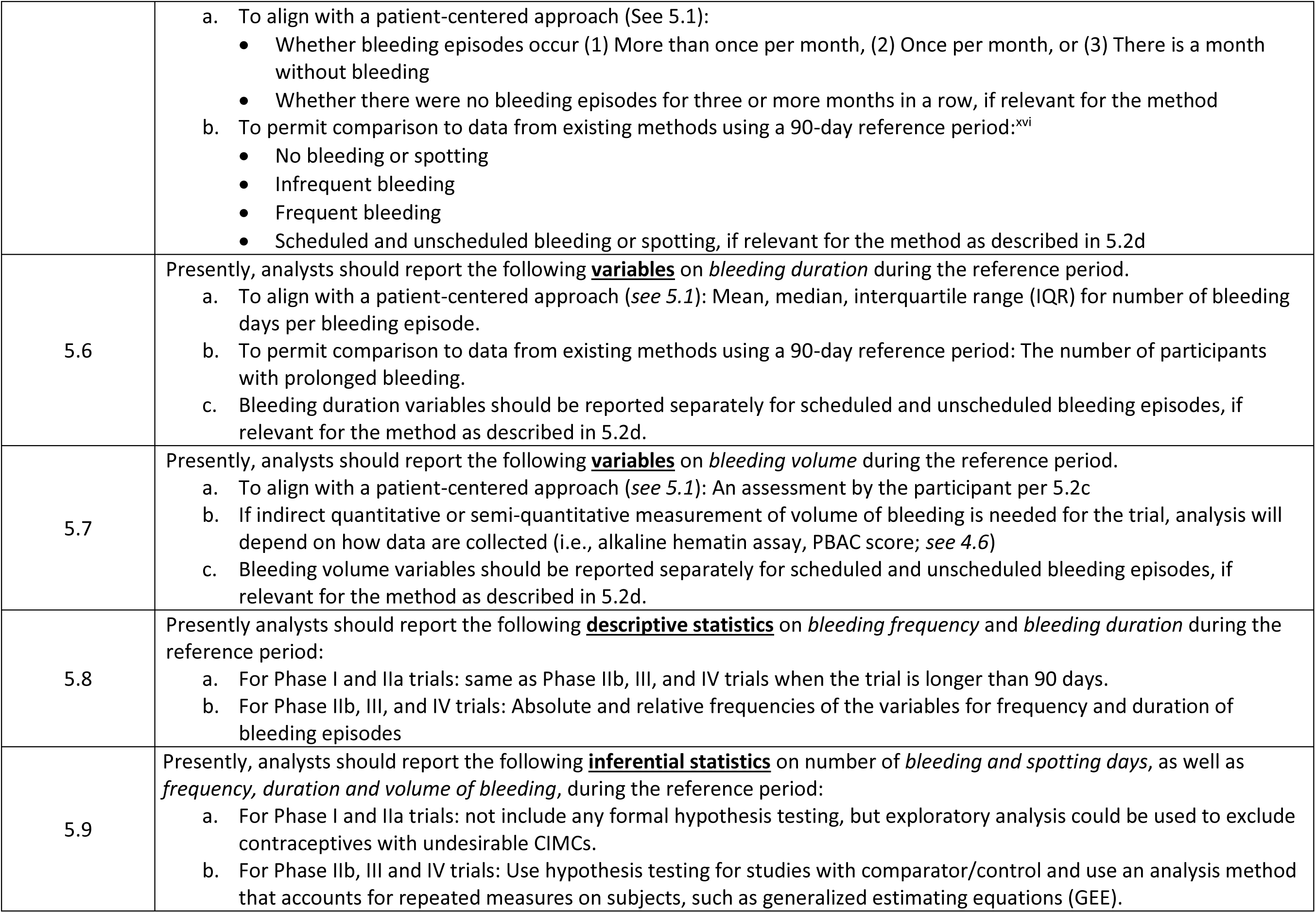
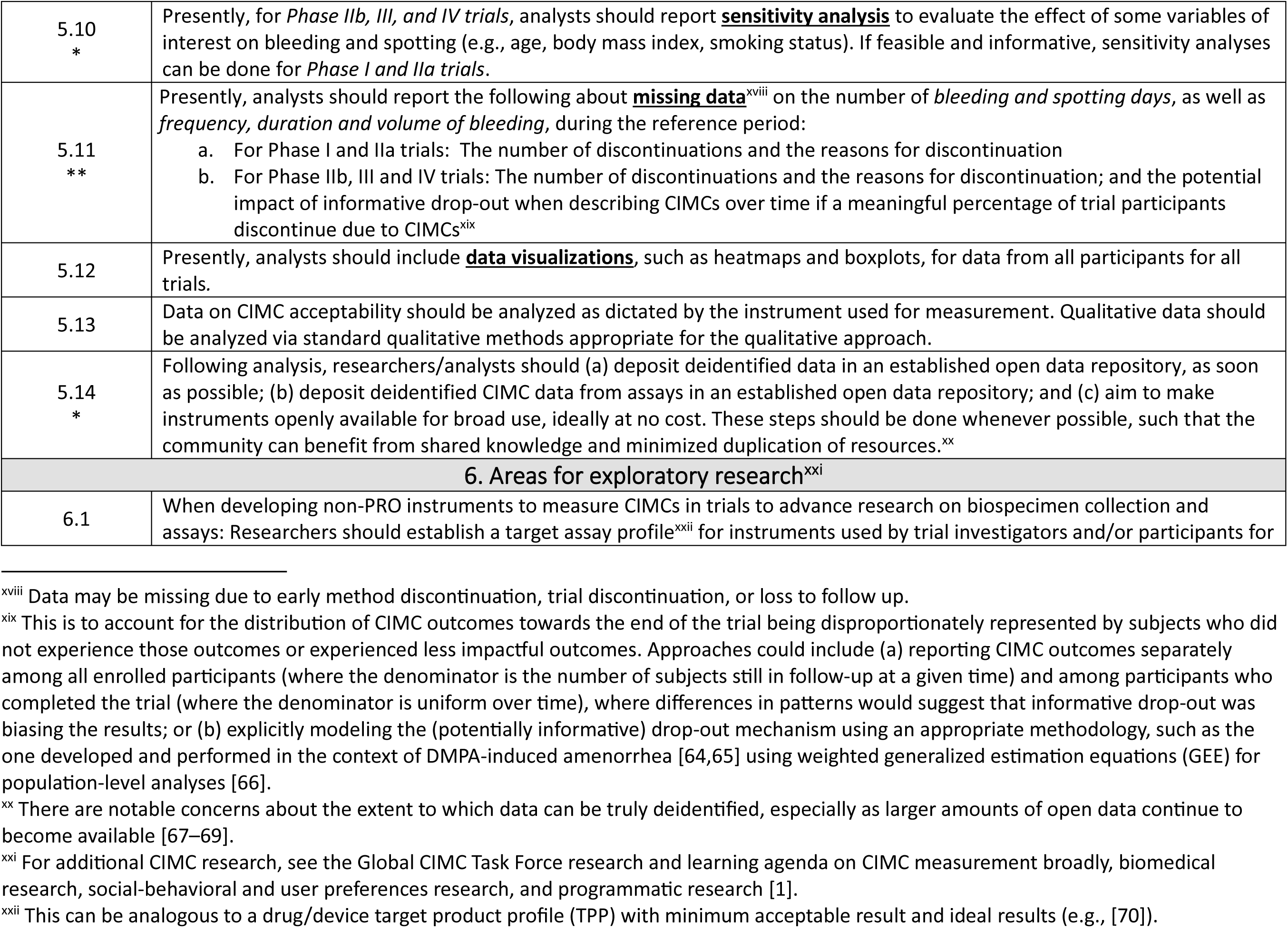
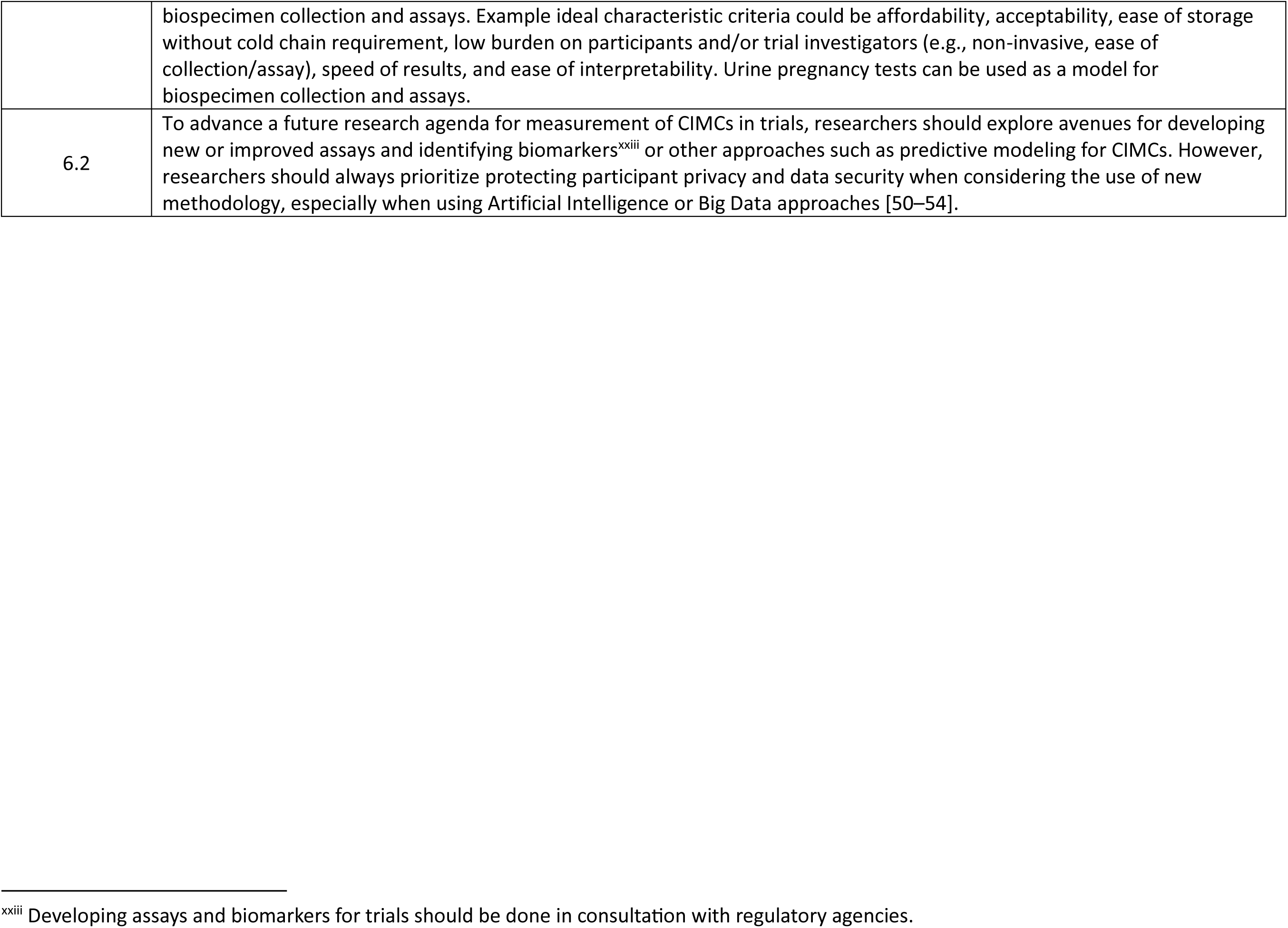
Consensus^i^ recommendations on CIMC measurement in contraceptive clinical trials.

|  |  |
| --- | --- |
| **\*\*** | >85% of experts agree that this recommendation should be prioritized (See Supplementary Table S3) |
| **\*** | 75%-84% of experts agree that this recommendation should be prioritized |
| **†** | Recommendation adapted from Creinin et al., 2022 [12] |
| **§** | Recommendation informed by regulatory guidance or best practices on measuring patient-reported outcomes (PROs), like CIMCs |

| Number | Recommendation |
| --- | --- |
| **1. Standardization recommendations** |  |
| 1.1<br>**\*\* §** | As much as possible, research on CIMCs should aim to:<br>a. Simplify terminology and/or align terminology with established standards to ease and improve global communication and collaboration around CIMCs in clinical trials among various stakeholders.<sup>ii</sup><br>b. Collect data and describe CIMCs in a standardized way to allow comparisons across studies and products; researchers may also include assessments of participants’ perceptions of and experience with CIMCs.<br>c. Use standardized simple, patient-centered terminology when reporting CIMC data. This terminology should be based on evidence and not be overly prescriptive. In addition, researchers and analysts should include previous analysis approaches and terminology when necessary to permit comparisons to data from previous studies of existing methods. |
| 1.2 | Researchers should ideally consider how any new instruments<sup>iii</sup> used to collect CIMC data relate to commonly used questions in previous trials of already approved contraceptive methods—even if those questions were not part of a validated instrument—to allow comparability of data between current contraceptive methods and new methods for providers, users, and researchers, including for future systematic reviews. *Also See 2.1 to 2.3.* |

<sup>i</sup> The consensus threshold was 75% agreement (i.e., responding “strongly agree” or “somewhat agree”), aligned with recommended Delphi methodology [30]. The level of consensus reached for each recommendation and agreement on priorities are reported in Supplementary Table S2.
<sup>ii</sup> These standards can include FIGO’s System 1 for nomenclature of normal and abnormal uterine bleeding [55], the Global CIMC Task Force’s definition of CIMCs [1], recommendations on bleeding data analyses in contraceptive studies [12], and other terminology harmonization efforts, especially those using methodology to establish consensus and priorities (e.g., Delphi method, Child Health and Nutrition Research Initiative [CHNRI] approach).
<sup>iii</sup> Definition of instrument: An approach or method to measure or assess the outcome of interest, including those that are: (a) participant/patient-reported, (b) researcher/clinician-reported, (c) observer-reported; (d) biomarker-based, or (e) assay-based, as well as both ‘objective’ or ‘subjective’ measurements that encompass both direct observations and/or perceptions (Adapted from [56])

|  |  |
| --- | --- |
| 1.3<br>* † | Additional research will be needed for alignment with recommendations 1.1, 1.2, and 2.1 to 2.3.<br>Presently—and until such research is complete—researchers should use the following standardized definitions for spotting and bleeding:<br>a. **Spotting:** Blood and other uterocervical fluid for which the person uses no menstrual products.<sup>iv</sup><br>b. **Bleeding:** Any amount of blood and other uterocervical fluid greater than spotting requiring menstrual product use.<br>Researchers should continue to study how diverse populations understand and differentiate spotting and light bleeding to generate more standardized language for measurement across trial populations. (*Also see 2.3b.*)<br>(*See 5.2 for additional definitions.*) |
| 1.4 | While it is important to keep CIMC measures consistent for comparability across trials, there should be some flexibility for local research teams to translate, adapt, and add to these tools based on the local culture and context. |
| 2. Instruments for measuring CIMCs |  |
| 2.1<br>* § | Researchers will need to develop new, standard instruments to measure CIMCs in trials because no validated instrument currently exists to address CIMC constructs of interest and for use in the context of contraceptive clinical trials [43]. Instruments for both (a) the menstrual changes themselves (e.g., changes in bleeding frequency, duration, volume; change in blood; changes in pain) and (b) the impact, acceptability<sup>v</sup>, and perceptions of those menstrual changes will be needed. Development of new, standard instruments can include: (a) generating and validating new items, and/or (b) modifying items from related, existing instruments used to measure changes to the menstrual cycle (e.g., instruments developed within the context of examining menstrual & gynecological disorders & symptoms). |

<sup>iv</sup> In some populations, the availability of menstrual products may be limited and thus reduce their use. Other populations may readily access menstrual products and use them differently than other populations would. When a trial is performed in such a population, the authors should describe this issue and how they choose to define spotting and bleeding for their particular population.

|  |  |
| --- | --- |
|  | Because additional research will be needed to develop and validate instruments in alignment with this recommendation, 4.1 to 4.13 describe data collection approaches recommended for trials conducted prior to this additional research and 5.1 to 5.13 describe related data analysis approaches.<br>Recommendations 2.2 and 2.3 provide additional details on instrument development and validation. |
| 2.2<br>** § | When developing instruments to measure CIMCs in trials, it is critical that researchers:<br>Follow current recommendations from regulatory agencies and professional societies and consortia, such as the FDA’s guidance on PRO<sup>vi</sup> use in trials [26] and the PROTEUS Consortium’s recommendations on measuring PROs effectively.<sup>vii</sup><br>Use simple, evidence-based patient-centered terminology both within instrument items, prioritizing the perspective of the contraceptive user and simplicity over the convenience of the researcher or imposing definitions or characterizations that may not be sufficiently objective to translate across contexts.<sup>viii</sup><br>Ensure instruments are accessible to those with differing literacy levels and people with disabilities, including consideration of pictorial instruments and adherence to current USG plain language guidelines [44] and Web Content Accessibility Guidelines (WCAG) [45] for digital content.<br>Ensure instruments adhere to data privacy requirements, legal protections, and ethical considerations to protect data and prevent use of data by any entity outside of those described in the trial informed consent forms. |
| 2.3<br>§ | When developing instruments to measure CIMCs in trials, ideally:<br>For participant-entered diary instruments, researchers should establish evidence<sup>ix</sup> on recall to determine the window of retrospective data entry permitted for each type of CIMC measured; such evidence may build upon previous research around heavy menstrual bleeding (HMB).<br>Researchers should establish the transferability<sup>x</sup> of instruments, including following recommendations such as the ISPOR principles on good practice for translation and cultural adaptation [46,47] because instruments are needed in different languages and sociocultural contexts, including for multisite trials. When feasible, this should include simultaneous development and validation of instruments in multiple contexts. (*Also see 1.3.*) |

<sup>vi</sup> All data on CIMCs in trials are reported by the participant (i.e., it is a patient reported outcome [PRO]) unless the trial is using an indirect quantitative and semi-quantitative approach like an alkaline hematin assay to determine menstrual blood loss from used menstrual products. See 4.6, 5.7b, 6.1, and 6.2 for recommendations on these types of instruments.
<sup>vii</sup> PROTEUS recommendations on measuring PROs effectively with ISOQOL standards [58].
<sup>viii</sup> If supported by evidence, an example of this could be asking participants about use/nonuse of menstrual products, instead of asking participants to use and understand the term ‘spotting’, which may not be relevant in all contexts.
<sup>ix</sup> “Establishing evidence” can include reviewing existing research and/or conducting new research.
<sup>x</sup> Transferability is the degree to which an instrument can be transferred between linguistic and sociocultural groups.

|  |  |
| --- | --- |
|  | c. Researchers should consider time and cost requirements to implement instruments, so they do not unduly hinder use in contexts with fewer resources.<br>d. For mixed mode instruments (e.g., paper and electronic instruments, or interactive voice response (IVR) and short message service (SMS)/chat instruments), researchers should establish equivalence<sup>xi</sup> between modes (e.g., between paper and electronic versions of an instrument), including following recommendation such as the ISPOR good research practices on use of mixed mode PROs [48].<br>e. For instruments requiring data entry by participants on electronic hand-held devices (i.e., phones or tablets), researchers should establish equivalence<sup>xi</sup> for devices provided to participants by the trial as well as for devices already used/owned by participants (i.e., BYOD, ‘bring your own device’) so either can be used by trials without the potential of introducing differential bias between trials providing devices and trials with BYOD or within trials where some participants are provided devices and some are BYOD [49].<br>f. Researchers should consider if additional instrument items or modules are needed for participants from populations who may have different CIMC outcomes and experiences (e.g., those with menstrual & gynecological disorders & symptoms). |
| **3. Trial design, protocol development, participant recruitment** |  |
| 3.1 | Early phase studies should consider evaluation of drug-drug interactions—including pharmacokinetic (PK) or pharmacodynamic (PD) interactions that may impact CIMCs—for commonly used medications that may influence CIMCs. These medications can include over the counter (OTC), supplements, or even certain foods. |
| 3.2<br>* | Every effort should be made to evaluate a range of body mass index (BMI)<sup>xii</sup> categories in PK studies, as BMI heterogeneity can have major implications for PK and/or CIMCs. |

<sup>xi</sup> Measurement equivalence is a function of the comparability of the psychometric properties of the data obtained via the original and adapted administration mode... to demonstrate that the change did not introduce response bias and/or negatively affect the measure’s psychometric properties” [48].

|  |  |
| --- | --- |
| 3.3<br>* | Related to intrinsic or extrinsic factors that may impact CIMCs:<br>a. Researchers should consider the most relevant intrinsic or extrinsic factors and design enrollment for Phase III and IV trials to include adequate numbers of participants with these factors. The choice of relevant factors will be informed by trial context, trial population, product characteristics, and/or trial design.<br>b. Information on intrinsic and extrinsic factors should be collected during the trial, as well as at enrollment.<br>c. When the primary trial outcome is pregnancy prevention, researchers should consider the impact of the selected intrinsic and extrinsic factors on risk of pregnancy.<sup>xiii</sup> |
| 3.4<br>§ | Researchers should establish and standardize what data are needed at enrollment on menstrual cycle history (when not using hormonal or intrauterine contraception) and other sexual and reproductive health history based on the CIMC data collected during the trial in order to permit comparisons between baseline and treatment outcomes. Additional baseline data on sociodemographics and other relevant background data may also be needed for stratified analyses to examine relevant intrinsic and extrinsic factors that may impact CIMCs. |
| 3.5 | When feasible, in Phase II and III trials, measurement of CIMCs and related outcomes should include input from local researchers, and whenever possible, also be under the advisement of local community advisory boards and community advocates. This input should incorporate input from a variety of stakeholders, including partners and families. |
| 3.6 | In Phase III and IV studies, diverse trial sites should be used to more fully represent differences in CIMC experiences and sociocultural differences in menstruation perceptions/knowledge across populations of future users.<sup>xiv</sup> |
| 3.7<br>* | Phase IV studies should be designed to build upon Phase III findings, to collect data on additional CIMC-related factors that may influence tolerability and method continuation, and to include adequate numbers of participants with these factors. The factors of interest may differ by trial context, product characteristics, and/or trial design. |
| 3.8 | All health care providers who will be involved in contraceptive clinical trials should be trained in comprehensive counseling methods, including providing information about different CIMCs and management strategies for CIMCs that are culturally and contextually appropriate and referring to appropriate services when indicated. |
| **4. Data collection** |  |
| 4.1<br>** | Additional research will be needed to align with recommendations 2.1 to 2.3. For trials conducted prior to this additional research, recommended data collection approaches are presented in 4.1 to 4.13 and recommended analysis approaches for these data are presented in 5.1 to 5.12.<br>Presently, researchers should include two types of assessments for primary outcomes of CIMCs:<br>Bleeding and spotting days, and<br>Frequency, duration, and volume of bleeding episodes. |
| 4.2<br>§ | Presently, researchers may also collect data on additional PROs such as cramping, pain, changes in blood and other uterocervical fluid, as well as other symptoms that have been shown to influence patient experiences with and perceptions of contraceptive methods. *See recommendations 4.11 to 4.13 for details.*<br>More research is needed to create validated instruments for these types of additional outcomes across trial contexts. *See 2.1 to 2.3 for details on instrument development and validation recommendations.* |
| 4.3<br>* | Presently, researchers should collect *bleeding and spotting data* in trial phases as follows:<br>In Phase I and IIa trials, researchers may choose to capture bleeding and spotting assessments as an exploratory outcome to make early decisions about product viability.<br>In Phases IIb and III, researchers should collect bleeding and spotting data.<br>In Phase IV, bleeding and spotting assessments may be required to assess additional use/benefits of the contraceptive (e.g., for special populations, health conditions, comparisons with other contraceptives). |
| 4.4<br>* | Presently, researchers should collect data on *frequency, duration, and volume* of bleeding episodes in trial phases as follows:<br>In phases IIb and III, these assessments are recommended.<br>In phase IV, these assessments may be required for additional use/benefits of the contraceptive (e.g., for special populations, health conditions, comparisons with other contraceptives). |
| 4.5 | Presently, researchers should collect some bleeding data for the entire duration of the method, as practical, including for methods with longer durations (e.g., 3-5 years). |
| 4.6<br>* | Presently, researchers should use validated measures for an objective measure of *bleeding volume* in trials when bleeding volume is an important outcome for label purposes (e.g., as an indication for HMB). |
| 4.7 | Presently, researchers should design bleeding data capture instruments to register information daily when possible and, if not, at least each time when bleeding or spotting occurs. However, researchers should study alternatives to daily collection of data for methods that have long durations (e.g., 3-5 years). |
|  | *(See 2.3a on recall for new instruments.)* |
| 4.8 | Presently, when using electronic diaries, the diaries should be designed to allow a window of time for users to retroactively record bleeding occurrences (e.g., within 48 hours); however, more research needs to be conducted to find the optimal window of time.<br>*(See 2.3a on recall for new instruments.)* |
| 4.9<br>* | Presently, researchers should consider the culture and context of their trial setting to determine whether paper or electronic diaries will be more acceptable and/or feasible and assess the strengths and limitations of each method of data collection.<br>*(See 2.3d on mixed modes for new instruments.)* |
| 4.10 | Presently, researchers using electronic diaries should attempt to incorporate data capture on a participant's own phone to avoid the burden of using additional devices where possible, despite software standardizing challenges.<br>*(See 2.3e on BYOD for new instruments.)* |
| 4.11<br>** § | Presently, trials should measure acceptability and quality of life related to CIMCs to assess the impacts of CIMCs on users and inform contraceptive counseling and user choices. This can be done in the following ways:<br>a. In Phase I, II, and III, researchers should measure participants' self-perceived changes in bleeding and other CIMCs (e.g., whether participants self-define changes as significant). This should be measured over time for Phase II and III.<br>b. In Phase II and III, researchers should measure the physical and psychosocial impacts of CIMCs on quality of life, including burdens and benefits. The aspects that are measured should be based on local priorities and informed by local researchers and advisors, and may include impacts on work, sexual well-being, mental burden, anxiety and stress, quality of relationships, social well-being, and financial well-being (including the burden of menstrual health management).<br>c. In Phase II and III, researchers should measure changes in acceptability related to CIMCs over time. |
| 4.12 | Presently, in Phase III, and when possible in Phase II, trials should include measures to contextualize and understand the CIMC acceptability and quality of life impacts outlined in recommendation 4.11. This can be done in the following ways:<br>a. Trials should collect sufficient demographic data, including about race and ethnicity, socioeconomic status, age, disability, sexual orientation, gender identity, location (e.g., urban/rural), and other social factors.<br>b. Trials should collect data about affordability and access to menstrual health information, menstrual products/materials, and water and facilities for changing, washing, and disposing of menstrual products.<br>c. Trials should collect data about the ethicality and cultural appropriateness of CIMCs (e.g., shame or stigma surrounding CIMCs) in the trial context, along with individuals' beliefs about CIMCs (e.g., the relationship between CIMCs and long-term health outcomes like infertility or cancer).<br>d. Researchers should measure self-efficacy related to managing CIMCs, that is, an individual's confidence in their capacity to manage the CIMCs experienced. |
| 4.13 | Presently, in all phases of trials, acceptability and impacts of CIMCs should be measured using both quantitative measures and qualitative methods whenever possible and advantageous. Qualitative methods will vary based on context and type of |
|  | trial but may include open-ended survey questions, in-depth interviews, or ranking exercises. Research teams should include members experienced in qualitative measurement and analysis whenever these methods are being used. |
| **5. Data analysis** |  |
| 5.1<br>† | Additional research will be needed to align with recommendations 1.1, 1.2 and 2.1 to 2.3, which will include establishing how new instrument(s) will be analyzed. For trials conducted prior to this additional research, recommended data analysis approaches are presented in 5.1 to 5.12 for data collected per 4.1 to 4.13.<br>Presently, to align with a patient-centered approach, data in the recommendations below should be reported for each 30-day **reference period**. To permit comparison to existing methods, data should also be reported for each 90-day reference period as well as other reference period length if relevant for the method.<sup>xv</sup> |
| 5.2<br>** † | Presently, researchers should use the following definitions during analysis for bleeding episode and for duration, frequency, and volume of bleeding episodes:<br>**Bleeding episode:** one or more consecutive days with bleeding and/or spotting, bordered by 2 full days without any bleeding or spotting.<br>**Duration:** A bleeding episode can be classified into prolonged or not prolonged.<br><u>Prolonged Duration</u> – Bleeding/spotting episode lasting more than 7 days.<br>**Volume:** Volume should be classified according to the participant’s perception. Volume should be described as “lighter,” “usual,” or “heavier” as compared to the individual’s typical volume when not using contraception.<br>**Frequency:**<sup>xvi</sup><br><u>Absence of bleeding/spotting:</u> no bleeding or spotting during the reference period.<br><u>Infrequent bleeding:</u> 2 or fewer bleeding/spotting episodes during a 90-day reference period.<br><u>Frequent bleeding:</u> > 4 bleeding/spotting episodes during a 90-day reference period. |

<sup>xv</sup> For example, 28 days for a combine oral contraceptive with 24/4 regimen (i.e., 24 days of pills with the APIs and 4 days of placebo pills) or other single cycle regimens (e.g., 21/7); or 84 days for an extended cycle combined oral contraceptive [12].

|  |  |
| --- | --- |
|  | • <u>Scheduled bleeding</u>: a bleeding/spotting episode starting during the expected interval and lasting no more than 7 days for a method expected to have a predictable bleeding pattern.<sup>xvii</sup><br>• <u>Unscheduled bleeding</u>: a bleeding/spotting episode occurring at any time prior to the expected interval, even if it continues into the next expected interval for a method expected to have a predictable bleeding pattern.<sup>xvii</sup> |
| 5.3<br>** | Presently, analysts should report the following **variables** on *number of bleeding and spotting days* during the reference period:<br>a. Total number of bleeding or spotting days,<br>b. Number of bleeding days,<br>c. Number of spotting days,<br>d. Number of scheduled and unscheduled bleeding or spotting days, if relevant for the method as described in 5.2d. |
| 5.4<br>* | Presently, analysts should report the following **descriptive statistics** on *number of bleeding and spotting days* during the reference period:<br>a. For Phase I and IIa trials, as exploratory summary statistics:<br>• Mean, median, interquartile range (IQR) for:<br>○ Bleeding or spotting days,<br>○ Bleeding days,<br>○ Spotting days,<br>○ Instances of consecutive days with no bleeding if daily bleeding data were collected, and<br>○ Scheduled and unscheduled bleeding or spotting days, if relevant for the method as described in 5.2d.<br>• Percent of participants with no bleeding or spotting.<br>b. For Phase IIb, III and IV trials:<br>• Mean, median, range, interquartile range (IQR),<br>• An estimate of precision (e.g., standard error of the mean or confidence interval)<br>• These data should be presented separately for total, unscheduled, and scheduled days if relevant for the method as described in 5.2d.<br>However, analysts are advised to consider whether the sample size and presumed distribution of the data warrants the use of standard statistical methods. |
| 5.5 | Presently, analysts should report the following **variables** on *bleeding frequency* during the reference period. |

<sup>xvii</sup> Predictable bleeding pattern: an expected predictable pattern is based on the product design as determined by the regimen (e.g., combined hormonal methods [oral, transdermal, vaginal, or injectable] and non-hormonal methods such as IUDs or non-hormonal vaginal products). Unpredictable bleeding pattern: an expected unpredictable pattern based on the product design (e.g., progestin only methods including implants, IUDs, injectables, pills, patches, and rings) [12].

|  |  |
| --- | --- |
|  | a. To align with a patient-centered approach (See 5.1):<br>• Whether bleeding episodes occur (1) More than once per month, (2) Once per month, or (3) There is a month without bleeding<br>• Whether there were no bleeding episodes for three or more months in a row, if relevant for the method<br>b. To permit comparison to data from existing methods using a 90-day reference period:<sup>xvi</sup><br>• No bleeding or spotting<br>• Infrequent bleeding<br>• Frequent bleeding<br>• Scheduled and unscheduled bleeding or spotting, if relevant for the method as described in 5.2d |
| 5.6 | Presently, analysts should report the following **variables** on *bleeding duration* during the reference period.<br>a. To align with a patient-centered approach (*see* 5.1): Mean, median, interquartile range (IQR) for number of bleeding days per bleeding episode.<br>b. To permit comparison to data from existing methods using a 90-day reference period: The number of participants with prolonged bleeding.<br>c. Bleeding duration variables should be reported separately for scheduled and unscheduled bleeding episodes, if relevant for the method as described in 5.2d. |
| 5.7 | Presently, analysts should report the following **variables** on *bleeding volume* during the reference period.<br>a. To align with a patient-centered approach (*see* 5.1): An assessment by the participant per 5.2c<br>b. If indirect quantitative or semi-quantitative measurement of volume of bleeding is needed for the trial, analysis will depend on how data are collected (i.e., alkaline hematin assay, PBAC score; *see* 4.6)<br>c. Bleeding volume variables should be reported separately for scheduled and unscheduled bleeding episodes, if relevant for the method as described in 5.2d. |
| 5.8 | Presently analysts should report the following **descriptive statistics** on *bleeding frequency* and *bleeding duration* during the reference period:<br>a. For Phase I and IIa trials: same as Phase IIb, III, and IV trials when the trial is longer than 90 days.<br>b. For Phase IIb, III, and IV trials: Absolute and relative frequencies of the variables for frequency and duration of bleeding episodes |
| 5.9 | Presently, analysts should report the following **inferential statistics** on number of *bleeding and spotting days*, as well as *frequency, duration and volume of bleeding*, during the reference period:<br>a. For Phase I and IIa trials: not include any formal hypothesis testing, but exploratory analysis could be used to exclude contraceptives with undesirable CIMCs.<br>b. For Phase IIb, III and IV trials: Use hypothesis testing for studies with comparator/control and use an analysis method that accounts for repeated measures on subjects, such as generalized estimating equations (GEE). |
| 5.10<br>* | Presently, for *Phase IIb, III, and IV trials*, analysts should report **sensitivity analysis** to evaluate the effect of some variables of interest on bleeding and spotting (e.g., age, body mass index, smoking status). If feasible and informative, sensitivity analyses can be done for *Phase I and IIa trials*. |
| 5.11<br>** | Presently, analysts should report the following about **missing data**<sup>xviii</sup> on the number of *bleeding and spotting days*, as well as *frequency, duration and volume of bleeding*, during the reference period:<br>a. For Phase I and IIa trials: The number of discontinuations and the reasons for discontinuation<br>b. For Phase IIb, III and IV trials: The number of discontinuations and the reasons for discontinuation; and the potential impact of informative drop-out when describing CIMCs over time if a meaningful percentage of trial participants discontinue due to CIMCs<sup>xix</sup> |
| 5.12 | Presently, analysts should include **data visualizations**, such as heatmaps and boxplots, for data from all participants for all trials. |
| 5.13 | Data on CIMC acceptability should be analyzed as dictated by the instrument used for measurement. Qualitative data should be analyzed via standard qualitative methods appropriate for the qualitative approach. |
| 5.14<br>* | Following analysis, researchers/analysts should (a) deposit deidentified data in an established open data repository, as soon as possible; (b) deposit deidentified CIMC data from assays in an established open data repository; and (c) aim to make instruments openly available for broad use, ideally at no cost. These steps should be done whenever possible, such that the community can benefit from shared knowledge and minimized duplication of resources.<sup>xx</sup> |
| **6. Areas for exploratory research<sup>xxi</sup>** |  |
| 6.1 | When developing non-PRO instruments to measure CIMCs in trials to advance research on biospecimen collection and assays: Researchers should establish a target assay profile<sup>xxii</sup> for instruments used by trial investigators and/or participants for |

<sup>xviii</sup> Data may be missing due to early method discontinuation, trial discontinuation, or loss to follow up.

|  |  |
| --- | --- |
|  | biospecimen collection and assays. Example ideal characteristic criteria could be affordability, acceptability, ease of storage without cold chain requirement, low burden on participants and/or trial investigators (e.g., non-invasive, ease of collection/assay), speed of results, and ease of interpretability. Urine pregnancy tests can be used as a model for biospecimen collection and assays. |
| 6.2 | To advance a future research agenda for measurement of CIMCs in trials, researchers should explore avenues for developing new or improved assays and identifying biomarkers<sup>xxiii</sup> or other approaches such as predictive modeling for CIMCs. However, researchers should always prioritize protecting participant privacy and data security when considering the use of new methodology, especially when using Artificial Intelligence or Big Data approaches [50–54]. |

<sup>xxiii</sup> Developing assays and biomarkers for trials should be done in consultation with regulatory agencies.

Our final recommendations include research approaches to establish the evidence for future improvement in CIMC measurement in trials (e.g., 2.1, 2.2 and 2.3) and recommendations investigators can implement in the meantime, while that research is being conducted (i.e., recommendations beginning with “presently…”, e.g., 4.1 through 4.13 and 5.2 through 5.12). As our aim was to build upon, and not replicate, previous work standardizing CIMC measurement in clinical trials, recommendations often incorporate concepts from previous criteria, especially the work of Creinin and colleagues [12].

Recommendations 1.3 and 5.2 draw heavily from these most recent criteria. Several recommendations also include and refer to regulatory guidance and best practices around developing measures for PROs like CIMCs.

Table 3. Consensus recommendations on CIMC measurement in contraceptive clinical trials See Table 3 <u>at end of document</u>.

## 4. DISCUSSION

Building upon previous approaches at standardization around CIMCs, our consensus recommendations were developed by virtually convening interdisciplinary experts from around the world to consider how to improve the current and future measurement and analysis of CIMC data in contraceptive clinical trials. The resulting 44 recommendations, including identified priorities, can serve as guidance to those implementing and funding clinical trials. Based on the thoughtful input throughout the expert consultation, there are a few important considerations we wish to highlight for those implementing our recommendations. We also note reflections in the Supplementary Material on our consensus-building methodology, which we propose as a model—for the field of contraception, wider sexual and reproductive health, and beyond—of how to successfully bring a large, interdisciplinary, and diverse group of people together from around the world to come to consensus on a topic that had eluded such previous agreement.

### 4.1 Reflections on developing recommendations

Although we ultimately reached consensus on almost every aspects of our recommendations, a few areas of contention emerged during the process. In some instances, differences occurred simply due to the interdisciplinary and global nature of our expert group and the distinct perspectives that inherently arise when convening those with different trainings, experiences, and areas of expertise. Often these differences emerged between those approaching the work from a biomedical perspective verses those working within social-behavioral research. All these differences and the ensuing conversations, however, were essential to thoroughly addressing a topic as multifaceted as measuring CIMCs in the context of clinical trials.

Some areas of contention proved especially challenging and often could be distilled down to differing perspectives along a continuum. On one end of the spectrum was a preference to recommend modest improvements to current approaches that would be feasible and straightforward to implement, aligning with past and present norms and existing systems; on the other end was a desire to push for larger, creative improvements aimed at innovating beyond current confines. In addition, we also encountered an innate tension in attempting to weigh the often-divergent perspectives of various stakeholders within the contraceptive clinical trial ecosystem, including trial investigators and other researchers, trial sponsors and funders, regulators, trial participants, and the future providers and users of the new contraceptives being developed in trials. Because we were developing recommendations for global use, balancing these different perspectives was even more challenging considering the wide range of contexts in which clinical trials are conducted, including within different healthcare systems, countries, and resource settings around the world. We describe a few examples of weighing these differing perspectives next.

Recommendations needed to be specific and technical enough to provide actionable guidance to trial investigators without being too burdensome to enact within the many constraints and competing demands of conducting a clinical trial. From this researcher perspective, there can be an appeal of smaller, incremental improvements to current practices that could be simple to implement with often-limited resources. This gradual approach could also be appealing from a sponsor and funder standpoint because it may not require increased trial budgets and may be more likely aligned with current and familiar regulatory strategies. For some experts, these types of modest changes within current norms and systems were of interest for very practical, realistic reasons founded in their many years of experience. Indeed, complex and/or numerous novel recommendations that may demand additional time and funding could result in delaying upcoming trials, as well as fewer, longer trials in the future. In addition, many experts were concerned our recommendations would be interpreted by funders or regulators as expert-endorsed mandates that should be required for every trial. Altogether, the broader impact of these types of additional burdens and constraints could be fewer new methods being developed, which could ultimately mean the contraceptive needs of some people and couples may not be met. To address the concerns our recommendations could hinder contraceptive R&D, we specify they should not be considered required for contraceptive trials to be funded, conducted, or reviewed by regulatory authorities. Rather, stakeholders should consider these as recommended approaches to achieving standardization and comparability across trials and to guiding future research for improvement, all with emphasis on identified priorities.

On the other hand, this convening of around 50 interdisciplinary experts from multiple global regions in a consensus-building process was a unique opportunity to push the contraceptive clinical trial ecosystem to consider innovative CIMC data collection and analysis approaches that could be more informative to providers in counseling on new contraceptives and more relevant to future users. Many experts endorsed this perspective, which is also aligned with current patient-focused drug development initiatives, other efforts at increasing patient engagement in clinical trials and regulatory decisions, and wider patient-centered outcome research [24–27,36,37]. As described in our recommendations, research will be needed to align CIMC measurement with best practices for PROs. Because this approach is novel for contraceptive trials—although increasingly common in other therapeutic areas—it will require close adherence to regulatory guidelines and engagement with regulators.

Although areas of contention persisted, our approach sought to balance the perspectives of all experts in the consensus-building process and stakeholders in the contraceptive clinical trial ecosystem in our final recommendations by focusing on the goal of improving the standardization and value of trial data on CIMCs. For example, this is the first time recommendations have included uterine cramping and pain, an important outcome for people who menstruate. By considering all CIMCs, not just changes in bleeding patterns, and including experts in menstrual health, heavy menstrual bleeding, patient-reported outcomes, social-behavioral researchers, and others, we were able to extend the boundaries of this work. One related limitation to our process we must acknowledge is the lack of direct input from clinical trial participants. Although we aimed to include their perspectives, as understood among our expert group, this does not replace explicit contributions from participants and their communities. To ensure the utility and relevance of the recommendations around future research a balanced and inclusive approach will need to continue, including using participatory-engaged research methodologies.

### 4.2 Implementing recommendations

As a starting point for implementing these recommendations, we have highlighted seven priorities: 1.1, 2.2, 4.1, 4.11, 5.2, 5.3, and 5.11 (listed separately in Supplementary Table S3). Recommendations 1.1, 2.2, and 5.2 also reached 100% consensus. Recommendation 1.1 calls for standardization and simplification of terminology. In support of this, 2.2 outlines best practices for making both terminology and measurement instruments simple, accessible, and patient-centered. Recommendation 5.2, also aligning with 1.1, lays out important terminology and definitions for use across trials, defining a bleeding episode, as well as duration, frequency, and volume of bleeding episodes. Recommendations 1.1 also calls for standardization in the type of CIMC data collected; therefore, 4.1 and 4.11 provide more detail on data collection, with 4.1 covering the primary outcomes of CIMCs that should be collected, including bleeding and spotting days and frequency, duration, and volume of bleeding episodes, and 4.11 outlining the acceptability and quality of life data to be collected. Finally, 5.3 and 5.11 outline some key variables and missing data that we recommend be reported for all contraceptive clinical trials.

Even implementing only these seven priority recommendations will come with challenges. We recognize the list of all recommendations in Table 3 is not particularly accessible or user-friendly to all. An important next step is to develop tools for stakeholders across the contraceptive R&D ecosystem, such as checklists and guidelines to make adopting these recommendations more straight-forward without imposing any unnecessary burden on the development of new contraceptives. It will also be valuable to tailor these resources to the types of research being conducted (e.g., Phase 1 vs. Phase 3, or clinical vs. social-behavioral research) and to target audiences (e.g., researchers, funders, providers, regulators).

### 4.3 Future implications

We call on stakeholders in the contraceptive clinical trial ecosystem to view these recommendations as new best practices, developed and endorsed by a wide range of experts in contraceptive R&D and related disciplines. We encourage those involved in the design of clinical trials to begin to incorporate these recommendations into their work as soon as possible. To achieve the full benefits of these recommendations, additional dissemination will be required. This publication is only the beginning of efforts needed to make these recommendations actual best practices amongst the contraceptive R&D community.

We also note contraceptives are not the only drug, device, or biologic to impact the menstrual cycle, yet such data are not routinely collected during clinical trials or during standard toxicology and pharmacodynamics studies, as other organ functions and vital signs are. The lack of these data was recently highlighted with the initial introduction of COVID vaccinations when vaccinated people who could or did menstruate experienced unanticipated bleeding changes [38–42]. We hope our work can be a guide and an invitation to develop recommendations for if, when, and how to measures changes to the menstrual cycle in more types of clinical trials and related research.

Broadly, our consensus recommendations can enable more standardized and patient-centered measurement and analysis of CIMC data in the near-term and continued improvement in the future, which can result in more accurate information in contraceptive product labeling on data that matter more to users. Ultimately, such efforts can allow for greater comparability across trials and better data synthesis in systematic reviews and meta-analyses that can inform clinical guidance. In turn, these improvements can enable providers to offer better counseling and users to make more informed decisions about their choice of method, which can improve the experiences of contraceptive users and better meet the contraceptive needs of people and couples.

### Supplementary material

1. Supplementary Material file
2. Supplementary tables

## Data Availability

All data produced in the present work are contained in the manuscript.

## Acknowledgments

This expert consultation was made possible by the generous support of the American people through the U.S. Agency for International Development (USAID), provided to FHI 360 through cooperative agreement 7200AA20CA00016. The contents are the responsibility of FHI 360, the views expressed are those of the authors, and contents and views do not necessarily reflect those of any funder, including USAID, the National Institutes of Health, the United States Government, or the Reckitt Global Hygiene Institute Fellowship. ACLM, SC, EH, NM, AEB, RLC, AFC, ECC, LJD, AM, KN, MS, DT, and EET were supported but USAID cooperative agreement 7200AA20CA00016, with SC also supported by the Bill & Melinda Gates Foundation (OPP1200867) and the Eunice Kennedy Shriver National Institute of Child Health and Human Development (T32 HD52468) and MS also supported by the Bill & Melinda Gates Foundation (INV-045483). EC was supported by Exeltis, and CEN by USAID. LB, VB, KBHC, MDC, HODC, GFD, AE, LBH, JH, CRTJ, SPSK, DM, KAM, JAM, FMO, CBP, CSV, RSW, JAS, LMS, and OV received an honorarium supported by USAID cooperative agreement 7200AA20CA00016 that was declined by other authors. In addition, GFD was also supported by USAID Cooperative Agreement 7200AA20CA00019 provided to CONRAD/Eastern Virginia Medical School, JH was also supported by a Reckitt Global Hygiene Institute Fellowship, JAM was also supported by Wellcome Fellowship 209589/Z/17/Z, FMO was also supported by EVIHDAF, and RSW was also supported by The Population Council.

The FHI 360 team would like to recognize the additional contributions of the external planning committee members, whose feedback greatly improved the consensus-building process. These members are Amanda Cordova-Gomez, Anita Nelson, Bellington Vwalika, Carolina Sales Vieira, Chelsea Polis, Diana Mansour, Funmi OlaOlorun, Kathryn Clancy, Kristen Matteson, Lisa Haddad, Mitchell Creinin, Olivia Vandeputte, and Tanya Dargan Mahajan. We also appreciate the *ad hoc* advisory roles of Lisa Rarick on regulatory considerations, Marni Sommer on menstrual health, and Greg Kopf on preclinical and early pharmaceutical trials. We thank Leigh Wynne for input on virtual meeting engagement approaches and coordinating Working Group 5. We also appreciate feedback from Aurélie Brunie and Holly Burke on the manuscript. Natt Pladsri and Audrey Fratus provided important support in preparing for the virtual meetings and ensuring they ran smoothly to aid in successful discussion.

