## Supplementary File for "Consensus recommendations for measuring the impact of contraception on the menstrual cycle in contraceptive clinical trials"

#### Consensus-building methodology

This supplementary file details our consensus-building methodology, which we propose as a model—for the field of contraception, wider sexual and reproductive health, and beyond—of how to successfully bring a large, interdisciplinary, and diverse group of people together from around the world to come to consensus on a topic that had eluded such previous agreement.

#### 1. Development

In September 2022, members of a scientific and technical team at [author organization] engaged a group of interdisciplinary experts about their interest in and availability for participating in a consensus-building process, their interest in joining a planning committee for that expert consultation, and their suggestions of other experts who should be included in the process. Invitees had a range of expertise in topics (i.e., contraceptive clinical trials, family planning, the menstrual cycle, menstrual health, measurement and analysis approaches, and regulatory considerations) and disciplines (i.e., clinical research, social-behavioral science, biomedical research, basic science, and clinical practice). Experts were recruited from members of the existing CIMC Global Task Force [1], contributors to previous standardization criteria, contributors to the literature in relevant fields, and the suggestions from other experts.

##### 1.1 External planning committee

We convened a planning committee of 13 external experts from 12 organizations and 6 countries to work with the [author organization] team to refine and improve the consensus-building process, to expand the list of invitees to include experts from more global regions, and to finalize the agenda for the first two days of meetings. We held three planning meetings in September, October, and December 2022.

First, the committee agreed upon three objectives for the expert consultation: (a) to review and discuss limitations and strengths of current approaches for measuring and analyzing data on CIMCs in contraceptive clinical trials; (b) to come to a consensus on recommendations for improvements in CIMC measurement and analysis in contraceptive clinical trials that meet the needs of trial participants, researchers, sponsors, regulators, and future users; and (c) to identify important, related topics outside the scope of this expert consultation that warrant similar consideration. The second objective also included determining what research and evidence are needed to empirically evaluate the recommended improvements and publication on the consensus-building process and recommendations.

Second, the [author organization] team developed an initial consensus-building methodology to propose to the planning committee based on the agreed-upon priorities that the process should: (a) allow for completely virtual participation, (b) not be too burdensome on participants, and (c) incorporate a mix of independent decision-making with debate and discussion. Because no single established approach met all these needs, the [author organization] team proposed an amalgamation of the RAND/UCLA method, the Delphi method, the Modified Rand-Delphi method, the Nominal Group Technique, and the Jandhyala method [2–7]. The planning committee suggested expanding the expert consultation from a proposed single convening over two half-days into a multi-month process. This expansion incorporated flexibility into the process so it could be modified or extended if needed and provided additional opportunities for asynchronous engagement. Additionally, the planning committee recommended hiring experienced external facilitators to support building consensus.

#### 1.2 Expert consultation process

The expert consultation consisted of a fully virtual consensus-building process including four half-day meetings, recommendation development within working groups, and a series of consensus questionnaires. By design, some details about the process were determined via group consensus and others were flexible to accommodate any needed modifications (e.g., the exact number of questionnaires would depend on the level of consensus reached in previous meetings and questionnaires); therefore, we present a summary of the initial process developed with the planning committee in *Supplementary File Figure 1* and details on the final process below in Results. Throughout the process, however, the following were consistent: (a) each of the four half-day meetings were scheduled during times within normal working hours for the most experts as possible and on dates when the most experts indicated they were available via Doodle polls; (b) all meetings took place on Zoom, with interactive features like real-time voting via the Zoom polling function to glean consensus, Google Jamboards to collect ideas and feedback, and/or Zoom breakout rooms for focused discussion and teambuilding; (3) each working group decided on the process for drafting and revising their own recommendations that best aligned with their schedules and preferred styles; (c) all consensus questionnaires asked about agreement/disagreement on a four-point Likert-like scale (i.e., strongly agree, somewhat agree, somewhat disagree, strongly disagree) and included at least one open-ended question; (d) primary qualitative analysis of open-ended questions was completed by one author (SC) with input from the [author organization] team; and (e) all documents used during the process, such as meeting materials, questionnaire results, collective resources, and working group documents, were housed in shared Google Drive folders to which all experts had access. The [Ethics Office] at [author organization] determined the expert consultation protocol was research that was exempt from Institutional Review Board review.

**Supplementary File Figure 1**. Summary of original consensus-building process for developing recommendations on CIMC measurement in contraceptive clinical trials resulting from planning committee and presented during the Day 1 meeting

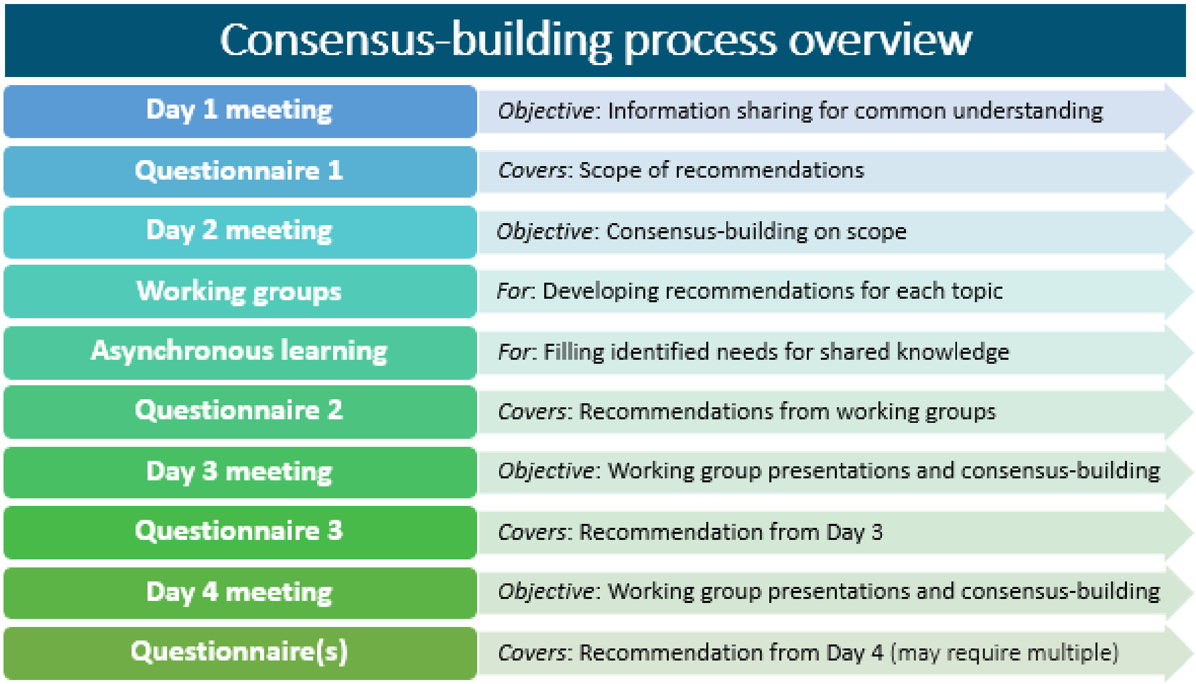


### 2. Outcomes

#### 2.1 Expert consultation process

We present the final process, reflecting consensus and iteratively developed decisions in *Figure 3* of the main paper. The following sections provide details on the process and results from each of three stages, which are color-indicated in *Supplementary File Figure 2*.

**Supplementary File Figure 2**. Final consensus-building process for developing recommendations on CIMC measurement in contraceptive clinical trials

| STEP | DETAILS |
|---|---|
| Day 1 meeting | *Objective*: Information sharing for common understanding |
| Questionnaire 1 (Q1) | *Covers*: Scope of recommendations |
| Day 2 meeting | *Objective*: Consensus-building on scope (i.e., topics included) |
| Working groups | *For*: Developing recommendations for each topic |
| Asynchronous learning | *For*: Filling knowledge gaps identified in Q1 and during meetings |
| Questionnaire 2 (Q2) | *Covers*: Recommendation drafted by Working Groups 1-3 |
| Sync-Up Call | *Covers:* Revisions to recommendations from Working Groups 1-3 |
| Questionnaire 3 (Q3) | *Covers*: Recommendation from all working groups |
| Day 3 meeting | *For*: Working group presentations and consensus-building |
| Day 4 meeting | *For*: Working group presentations and consensus-building |
| Finalize Recommendations | *Objective*: Compile, streamline, and organize recommendations |
| Prioritization Survey | *Covers:* Final recommendations and prioritization |

| Knowledge Sharing and Defining Scope | Formation of Recommendations by Working Groups | Final Consensus-Building for Recommendations |
|---|---|---|

*Notes: Members of the [author organization] team who developed Questionnaire 1 were not eligible to respond to it because their input was already incorporated; however, they were eligible to respond to all others. The five working groups were on eligibility, data, acceptability, instruments, and analysis (Supplementary File Table 1).*

#### 2.2 Knowledge sharing and defining the scope

The **Day 1** virtual half-day meeting took place on January 10, 2023. Our primary goal was establishing a common understanding of key information amongst our interdisciplinary group. The meeting consisted of interactive breakout sessions to establish or strengthen relationships between all experts, an overview of the consensus-building process, and presentations on key information needed to ensure a shared knowledge base. *Supplementary File Figure 3* shows the Day 1 meeting agenda and details about the presentations on contraceptive clinical trials, patient-reported outcome measures, sociocultural contexts of CIMCs, and inclusive language around gender and sexuality. Throughout the expert consultation process, information and resources on additional topics that warranted a similar common understanding, as well as updates on Day 1 topics, were shared with experts for asynchronous learning or shared via working groups.

**Supplementary File Figure 3**. Day 1 meeting agenda for consensus-building process to develop recommendations on CIMC measurement in contraceptive clinical trials

| EXPERT CONSULTATION DAY 1 AGENDA<br>TUESDAY, JANUARY 10, 2022 | | |
|---|---|---|
| 8:30-8:45 | **Welcome & Introduction to Consultation** | *Co-Facilitators: [names]* |
| 8:45-9:00 | **Breakout Session 1** | *All* |

| | | |
|---|---|---|
| 9:00-10:10 | **Session 1 – Technical Essentials**<br>• Contraceptive clinical trial essentials<br>• Patient-reported outcome measures (PROMs) in clinical trials<br>• Recommendations for standardization of bleeding data analyses<br>• Findings from menstrual changes measurement systematic review<br>Q&A | *Moderator: [name], USAID*<br>• *[name], Population Council*<br>• *[name], PROTEUS Consortium*<br>• *[name], UC Davis*<br>• *[name], FHI 360* |
| 10:10-10:25 | **Break** | *All* |
| 10:25-10:40 | **Breakout Session 2** | *All* |
| 10:40-11:40 | **Session 2 – Context and Inclusivity**<br>• Sociocultural context surrounding CIMCs<br>• Sharing the voice of experiences with CIMCs<br>• Inclusive language around gender and sexual activity<br>Q&A | *Moderator: [name], Univ of Ibadan*<br>• *[name], UNAIDS*<br>• *Anonymous stories*<br>• *[name], NIH* |
| 11:40-11:55 | **Breakout Session 3** | *All* |
| 11:55-12:05 | **Day 2 Preparations**<br>• Overview of Day 2 process<br>• Ground rules brainstorming | *Co-facilitators* |
| 12:05-12:30 | **Questionnaire 1 and Doodle Poll** | *All* |

At the end of the Day 1 meeting, experts received the first questionnaire (**Questionnaire 1**) to collectively decide which topics would be in or out of scope for the expert consultation. Topics proposed in Questionnaire 1 were developed by the [author organization] team with input from the planning committee. In Questionnaire 1, experts also had the opportunity to suggest additional topics for consideration by the group via voting during the Day 2 meeting. The **Day 2** virtual half-day meeting was on January 12, 2023, during which we agreed on ground rules for the consensus-building and finalized the scope of the recommendations. See *Supplementary File Figure 4* for the Day 2 meeting agenda.

**Supplementary File Figure 4**. Day 2 meeting agenda for consensus-building process to develop recommendations on CIMC measurement in contraceptive clinical trials

| **EXPERT CONSULTATION DAY 2 AGENDA<br>THURSDAY, JANUARY 12, 2022** | | |
|---|---|---|
| 8:30-8:55 | **Welcome & Process Review** | *Co-facilitators: [names]* |
| 8:55-9:35 | **Q1: Qualitative Feedback** | *Co-facilitators* |
| 9:35-10:00 | **Q1: Quantitative Feedback & New Topics** | *Co-facilitators* |

| | | |
|---|---|---|
| 10:00-10:15 | **Break** | *All* |
| 10:15-11:30 | **Working group breakout session** | *Co-facilitators* |
| 11:30-11:45 | **Plenary report-out on working group plans** | *All* |
| 11:45-12:30 | **Final Questions & Wrap-up** | *Co-facilitators* |

We report the recommendation scope agreed upon during Questionnaire 1 and Day 2 in *Table 2* of the main paper. Before Questionnaire 1, we had decided a topic would need 85% consensus (i.e., 'strongly agree' or 'somewhat agree') to be considered within the scope of our work, and all 11 topics proposed in Questionnaire 1 reached consensus for inclusion. Experts suggested another five topics, and although none reached consensus during voting on Day 2, a number of elements of those topics were incorporated into other consensus topics. Topics determined to be out of scope via Questionnaire 1 and Day 2 and during later stages of the consensus-building process are presented in *Supplementary Table S1* in the Supplementary Tables file, aligning with the third objective of the expert consultation to identify important, related topics that warrant similar consideration outside of the present process.

#### 2.3 Developing recommendations by Working Groups

After defining the scope of our recommendations, Day 2 also included forming working groups based on those topics. We condensed consensus topics into five areas around which working groups would develop recommendations (*Table 2* of the main paper). Experts self-selected into the five working groups according to their expertise and interest. A few experts, rather than joining a working group, opted to serve as *ad hoc* advisors for their areas of expertise to provide input, as needed, to the working groups.

We report on the members and scope of the five working groups in *Supplementary File Table 1*. Working groups included: (a) eligibility & enrollment requirements and confounders (i.e., Eligibility, for short), (b) type, frequency, and format/mode of CIMC data collected (i.e., Data), (c) measurement of acceptability & impact on daily life and culture & context (i.e., Acceptability), (d) a research agenda for developing CIMC instruments (i.e., Instruments)[i], and (e) analysis methodology and measure & analysis standardization (i.e., Analysis). From the consensus topics, we also identified three cross-cutting themes—(a) regulatory considerations; (b) incorporating user perspectives and understandings of CIMCs; and (c) culture and context—which would be considered across the recommendations from all working groups.

**Supplementary File Table 1**. Topical working group scope and membership for developing recommendations on CIMC measurement in contraceptive clinical trials

| Working Group | Scope | Members |
|---|---|---|

[i] During the expert consultation process, some members of the [author organization] team conducted a systematic review of how changes to the menstrual cycle have been measured using validated instruments across any discipline and caused by any etiology [9]. Updates on these review results were presented throughout the process to inform recommendations from the Instrument and other working groups.

| | | |
|---|---|---|
| 1. Eligibility | • Eligibility & enrollment requirements<br>• Confounders | [name], [name]*, [name], [name], [name], [name], [name], [name] |
| 2. Data | • Type, frequency, and format/mode of CIMC data collected | [name], [name], [name], [name], [name]*, [name], [name], [name] |
| 3. Acceptability | • Measurement of acceptability & impact on daily life<br>• Culture & context | [name], [name], [name], [name], [name]*, [name], [name], [name], [name], [name], [name], [name] |
| 4. Instruments | • Research agenda for developing CIMC instruments | [name], [name], [name], [name], [name]*, [name], [name], [name], [name] |
| 5. Analysis | • Analysis methodology<br>• Measures & analyses standardization | [name], [name], [name], [name], [name], [name]*, [name], [name], [name] |
| **Cross-cutting themes across Working Groups** | 1. Regulatory considerations<br>2. Incorporating user perspectives & understandings of CIMCs<br>3. Culture & context† | |
| ***Ad hoc* advisors across Working Groups** | [name] (regulatory), [name] (preclinical and early pharmaceutical trials), and [name] (menstrual health) | |

**Working group chair(s)*
*† Culture & context were both under the purview of Working Group 3 and a cross-cutting theme for all working groups to consider in their recommendations.*

Working groups convened between January and May 2023. Each working group developed their recommendations via multiple virtual calls and providing asynchronous input via document editing and/or email; some working groups used surveys to refine or prioritize their recommendations. During Day 2, we had decided recommendations would have to reach 75% agreement (i.e., ‘strongly agree’ or ‘somewhat agree’) by the expert group to be considered to have reached consensus, in line with recommended Delphi methodology [8]. We also decided on a sequential process for developing recommendations because recommendations created by some working groups would partially dictate those of other working groups (e.g., what data are collected per the Data Working Group would impact how that data are analyzed per the Analysis Working Group).

There were, therefore, two phases to this sequential process. In Phase one, the Eligibility, Data, and Acceptability Working Groups drafted their recommendations and received feedback from the wider group on those recommendations through **Questionnaire 2**. During this time, the Instruments and Analysis Working Groups also reviewed resources and discussed approaches for their recommendations. To bridge any gaps between working groups during Phase one, one author (AM) joined all working group meetings and emails to facilitate sharing across all groups.

In Phase two, the Instruments and Analysis Working Groups drafted their recommendations while the Eligibility, Data, and Acceptability Working Groups revised their recommendations. All working groups received feedback from the wider group on their drafted or revised recommendations through

**Questionnaire 3**. We also held a "sync up" call partway through Phase Two for working groups to share their progress, and shared an optional two-question survey to hear from experts on what they were most excited about from this work and any concerns they had in order to capture additional input and feedback during the process.

#### 2.4 Final consensus-building for recommendations

The **Day 3** virtual half-day meeting took place on May 16, 2023. During the meeting, each Working Group briefly presented their recommendations that had reached the 75% consensus threshold, and then reviewed recommendations that had not met consensus, including qualitative feedback from Questionnaires 2 and 3. For each Working Group, experts then had the opportunity to provide suggestions or additional reflections on recommendations that had not reached consensus and to discuss if there were any areas around which recommendations were missing. Working Groups met in Zoom breakout rooms for immediate discussion of the feedback received in the larger group session for recommendation revisions. In the week between the Day 3 and Day 4 meetings, the Working Groups reconvened, as necessary, to review and revise the recommendations that did not reach consensus, continue to review qualitative feedback from Questionnaire 3, and identify any recommendations that met consensus but also had relevant qualitative feedback that could be incorporated into a revision. The **Day 4** meeting was on May 25, 2023, during which Working Groups presented revised recommendations for a live vote via polling in Zoom. We also used this voting procedure to approve recommendations that Working Groups proposed to delete rather than revise. For recommendations that still did not reach 75% agreement, experts could then contribute to an open discussion to provide the Working Group with additional feedback, and Working Groups met in breakout rooms to further revise recommendations. In some cases, we edited recommendations live during this discussion and held additional rounds of voting. See *Supplementary File Figure 5* for Day 3 and Day 4 meeting agendas.

**Supplementary File Figure 5**. Days 3 and 4 meeting agenda for consensus-building process to develop recommendations on CIMC measurement in contraceptive clinical trials

| **EXPERT CONSULTATION DAY 3 AGENDA<br>TUESDAY, MAY 16, 2023** | | |
|---|---|---|
| **9:00 - 9:15** | Welcome & Objectives | *Co-facilitators: [names]* |
| **9:15 - 10:30** | Working Groups (WGs) 1, 2, 5 Presentations & Discussions | *WG Leads* |
| **10:30 - 10:45** | Break | *All* |
| **10:45 - 11:00** | Discussion about any missing recommendations | *Co-facilitators* |
| **11:00 - 11:50** | WGs 3, 4 Presentations & Discussions | *WG Leads* |
| **11:50 - 12:05** | Discussion about any missing recommendations | *Co-facilitators* |
| **12:05+** | WGs meet in breakout rooms to discuss updates to recommendations | *All* |

| | | |
|---|---|---|
| **5 minutes** | Close | *Co-facilitators* |

| **EXPERT CONSULTATION DAY 3 AGENDA**<br>**THURSDAY, MAY 25, 2023** | | |
|---|---|---|
| **10:00-10:10** | Welcome & Objectives | *Co-facilitators: [names]* |
| **10:10-10:30** | Working Group (WG) updates on revised recommendations from Day 3 | *WG Leads, WG Members* |
| **10:30-11:00** | Discussion of key themes and rollout of recommendations | *All* |
| **11:00-11:10** | Break | *All* |
| **11:00-11:40** | WG Breakout Sessions | *WG* |
| **11:40-11:55** | Break | *All* |
| **11:55-12:40** | Present and vote on revised recommendations | *All* |
| **12:40-1:00** | **Next steps & close** | **Co-facilitators** |

Following Day 4, the team at [author organization] **compiled and streamlined the recommendations** using all expert feedback to develop a full version of the recommendations for publication. Based on suggestions from experts during Questionnaire 3, Day 4, and elsewhere during the consensus-building process to this point, the compilation process included organizing recommendations into conceptual themes—sometimes beyond the topics of Working Groups, streamlining and combining similar recommendations to remove repetition, noting related recommendations, and minor revisions for clarity, alignment of terminology, and to reflect the new recommendation organization. There were five instances where all or part of a recommendation in the compiled list had not yet reached consensus, and most required at least some consensus for the recommendations to be usable and complete (e.g., an agreement on the definition of 'spotting'). For these recommendations, the [author organization] team drafted a revision based on feedback from the questionnaires and meetings for final consideration by the group.

During Day 4, we decided to add a **prioritization survey** based on the impact and urgency of the recommendation to aid trial implementors in identifying which should be considered a priority. Experts also had the opportunity to review the compiled, streamlined recommendations and provide any final feedback. In addition, experts were asked about the five revised recommendations that had not yet reached consensus to identify any final areas of agreement.

#### 3. Reflections on the consensus-building process

From the beginning, input from the external planning committee was critical to the success of consensus-building, and open feedback was important throughout the process. Experts did note the process was long and had the potential to be burdensome, especially with the need for consistency and standardization across the recommendations developed by different working groups. Overall, though, expert feedback was positive, with an appreciation for the flexible and iterative nature of the process and a general view that the consultation process was a valuable opportunity.

Our decision to organize a fully virtual convening was both a strength and a limitation. It was a priority for this work to bring together experts from all over the world in a series of meetings that permitted time for thoughtful discussion and consensus-building. Coordinating these meetings in-person over the five months would have been complex and prohibitively expensive, likely meaning we would have had to limit the amount of collaborative discussion time and/or restrict participation to experts from a smaller geographic area. Other similar efforts at standardization and consensus-building can often take place in-person at annual or biennial conferences of professional societies for a particular discipline or specialty; however, this was not possible for our interdisciplinary group, for whom there is no one—or even two to three—such meetings between which we could coordinate.

Virtual convenings, however, are not without faults. Although we included interactive and teambuilding elements into our virtual calls, this could not fully replicate the level of mutual understanding and collaboration that is possible in person. Additionally, our expert group was situation across 10 time zones, making scheduling the four half-day meetings impossible to do during regular working hours for all participants. Although we tried to include as much asynchronous work as possible outside of these meetings and always scheduled them at times for which the most experts would be within regular working hours and used Doodle polls, the meetings were likely easiest to attend for those in UTC-6 to UTC+3 time zones, which includes Africa, Europe, Latin America and the Caribbean, and the eastern part of Northern America, but not Asia and Oceania. Future work could explore ways of coordinating multiple, regionally-based convenings—either in-person or simply during regular working hours—who could then collaborate globally may be a way to address this limitation while increasing engagement across time zones and regions. In addition, for experts in any location with internet access limitations, active, full participation in the process would have been difficult.

Despite the size and differences within our expert group, consensus was reached in higher numbers and more often than initially expected. We propose this consensus-building process as a model—for the field of contraception, wider sexual and reproductive health, and beyond—of how to successfully bring a large, interdisciplinary, and diverse group of people together from around the world to come to consensus on a topic that had eluded such previous agreement. We also view this work as an example of how sexual and reproductive health research can and should embrace more patient-centered approaches, which is essential for our field as we continue to work towards better achieving the principles of informed choice, human rights, and justice.

#### 4. References

[1] Hoppes E, Nwachukwu C, Hennegan J, Blithe DL, Cordova-Gomez A, Critchley H, et al. Global research and learning agenda for building evidence on contraceptive-induced menstrual changes

for research, product development, policies, and programs. Gates Open Res 2022;6:49. https://doi.org/10.12688/gatesopenres.13609.1.

[9] Mackenzie A, Chung S, Hoppes E, Mickler A, Cartwright A. Measurement of changes to the menstrual cycle: a transdisciplinary systematic review evaluating measure quality and utility for clinical trials. Submitted to PloS One n.d.
