## Supplementary Tables S1-S3 for "Consensus recommendations for measuring the impact of contraception on the menstrual cycle in contraceptive clinical trials"

### Table S1. Identified topics outside the scope of the expert consultation process that may warrant similar consideration

| Topic | Discussion Details | Discussion about Scope |
|---|---|---|
| **CIMCs in pre-clinical development** | Debate about if the expert consultation scope should include pre-clinical development research such as use of animal models that menstruate, as well as organoids, physiomimetics, other *in vitro* organ systems. | Determined to be out-of-scope during the Day 2 meeting |
| **Expanding the definition of “acceptability” of contraception** | Discussion around how we generally define acceptability, and if we want to radically expand the definition with a higher bar to push acceptability beyond “minimally acceptable” | Determined to be out-of-scope in working group discussions, and WG will use existing definitions of acceptability. |
| **Balancing efficacy and acceptability data to support contraceptive counseling** | Role of expert consultation in advocating for contraceptive clinical trials to emphasize acceptability data in conjunction with efficacy data, specifically in an effort to support clinicians in counseling efforts. | Determined to be out-of-scope in working group discussions. |
| **Coherence** | Coherence (i.e., the extent to which the participant understands how the intervention works) as an important domain to measure in contraceptive clinical trials. | Determined to be out-of-scope in working group discussions. |
| **Sexual pleasure and well-being** | Discussion around the importance of including sex and pleasure and the positive | Determined to be out-of-scope in working group discussions. |

| | | |
|---|---|---|
| | | aspects of contraceptive use when measuring acceptability and preferences in clinical trials. |

**Table S2. Consensus agreement and priorities for recommendations on CIMC measurement in contraceptive clinical trials via Questionnaire 3 (Q3), Day 4 meeting (D4), and the Prioritization Survey (PS)**

| Rec. | Consensus | ≥90% consensus | 80-89% consensus | Priority agreement |
|---|---|---|---|---|
| 1.1 | a: 100% (Q3)<br>b: 92% (Q3)<br>c: 92% (Q3) | Yes | — | 100% |
| 1.2 | 100% (Q3) | Yes | — | 67% |
| 1.3 | a: 78% (PS)<br>b: 85% (Q3)<br>c: 100% (Q3) | No | No | 78% |
| 1.4 | 100% (Q3) | Yes | — | 61% |
| 2.1 | 100% (Q3) | Yes | — | 78% |
| 2.2 | a: 92% (Q3)<br>b: 100% (Q3)<br>c: 100% (Q3)<br>d: 100% (Q3) | Yes | — | 100% |
| 2.3 | a: 88% (Q3)<br>b: 96% (Q3)<br>c: 92% (Q3)<br>d: 95% (Q3)<br>e: 88% (Q3)<br>f: 96% (Q3) | No | Yes | 61% |
| 3.1 | 81% (Q3) | No | Yes | 28% |
| 3.2 | 96% (Q3) | Yes | — | 83% |
| 3.3 | a: 89% (D4)<br>b: 100% (Q3)<br>c: 94% (PS) | No | Yes | 78% |
| 3.4 | 100% (Q3) | Yes | — | 72% |
| 3.5 | 88% (Q3) | No | Yes | 56% |
| 3.6 | 88% (Q3) | No | Yes | 72% |
| 3.7 | 96% (Q3) | Yes | — | 83% |
| 3.8 | 77% (Q3) | No | No | 72% |
| 4.1 | 92% (Q3) | Yes | — | 94% |

| Rec. | Consensus | ≥90% consensus | 80-89% consensus | Priority agreement |
|---|---|---|---|---|
| 4.2 | 96% (Q3) | Yes | — | 56% |
| 4.3 | a: 85% (Q3)<br>b: 89% (Q3)<br>c: 89% (Q3) | No | Yes | 78% |
| 4.4 | 92% (D4) | Yes | — | 83% |
| 4.5 | 85% (Q3) | No | Yes | 67% |
| 4.6 | 94% (D4) | Yes | — | 78% |
| 4.7 | 92% (Q3) | Yes | — | 67% |
| 4.8 | 92% (Q3) | Yes | — | 72% |
| 4.9 | 100% (Q3) | Yes | — | 78% |
| 4.10 | 88% (Q3) | No | Yes | 56% |
| 4.11 | 3.1: 96% (Q3)<br>a: 96% (Q3)<br>b: 95% (D4)<br>c: 92% (Q3) | Yes | — | 89% |
| 4.12 | 3.2: 92% (Q3)<br>a: 92% (Q3)<br>b: 81% (Q3)<br>c: 81% (Q3)<br>d: 85% (Q3) | No | Yes | 61% |
| 4.13 | 92% (Q3) | Yes | — | 67% |
| 5.1 | 85% (Q3) | No | Yes | 61% |
| 5.2 | A: 100% (Q3)<br>b: 85% (Q3)<br>c: 85% (Q3)<br>d: 96%/89% (Q3)[i] | No | Yes | 100% |
| 5.3 | a. 92% (Q3)<br>b. 92% (Q3)<br>c. 85% (Q3)<br>d: 85% (Q3)<br>e: Not reached (PS)[ii] | No | No | 94% |
| 5.4 | a: 83% (PS)<br>b: 81% (D4) | No | Yes | 78% |
| 5.5 | a: 81-85% (Q3)[iii]<br>b: 81-89% (Q3)[iv] | No | Yes | 72% |

[i] When part of an earlier recommendation, 96% agreement was for all definitions related to ‘predictable’, and 89% agreement was for all related to ‘unpredictable’.

[ii] “Number of consecutive days with no bleeding for each instance of no bleeding if daily data were collected” did not reach consensus at 66% in the PS after further revision following Revision and discussion during D4.

[iii] 5.5a is a combination of two related parts of a recommendations, which both reached consensus during Q3, 81% for one and 85% for the other.

[iv] 5.5b is a combination of related parts of a recommendations, which all reached consensus during Q3 ranging from 81% to 89% (I.e., 89%, 81%, 85%, 85%).

| Rec. | Consensus | ≥90% consensus | 80-89% consensus | Priority agreement |
|---|---|---|---|---|
| 5.6 | a: 77% (Q3),<br>b: 85% (Q3[v])<br>c: 92% (Q3) | No | No | 56% |
| 5.7 | a: 81% (Q3)<br>b: 95% (D4)<br>c: 92% (Q3) | No | Yes | 56% |
| 5.8 | 91% (D4) | Yes | — | 72% |
| 5.9 | 88% (D4) | No | Yes | 56% |
| 5.10 | 100% (D4) | Yes | — | 78% |
| 5.11 | 77% (Q3) | No | No | 94% |
| 5.12 | 88% (Q3) | No | Yes | 44% |
| 5.13 | 95% (D4) | Yes | — | 67% |
| 5.14 | 92-96% (Q3)[vi] | Yes | — | 83% |
| 6.1 | 77% (Q3) | No | No | 33% |
| 6.2 | 79% (PS) | No | No | 56% |

Q3: Questionniare 3, D4: Day 4, PS: Prioritization survey

[v] The definition of prolonged bleeding differed between the Data and Analysis Working Groups, reflecting a difference between the International Federation of Gynecology and Obstetrics (FIGO) and the American College of Obstetricians and Gynecologists (ACOG). In Questionniare 3, "more than 8 days" (per FIGO) had 69% agreement, and "more than 7 days" had 85% agreement. To confirm this decision, experts completed a one-question survey about prolonged bleeding, and a majority agreed "more than 7 days" should be used for prolonged bleeding in all recommendations.

[vi] 5.14 is a combination of two related recommendations from different working groups, which both reached consensus during Q3, 92% for one and 96% for the other.

### Table S3. Priority Recommendations

<table>
<tr><th colspan="2">Recommendations with >85% of Agreement about Prioritization (n=7)</th></tr>
<tr><th>Number</th><th>Recommendation</th></tr>
<tr><td>1.1</td><td>As much as possible, research on CIMCs should aim to:<br>a. Simplify terminology and/or align terminology with established standards to ease and improve global communication and collaboration around CIMCs in clinical trials among various stakeholders.[vii]<br>b. Collect data and describe CIMCs in a standardized way to allow comparisons across studies and products; researchers may also include assessments of participants' perceptions of and experience with CIMCs.<br>c. Use standardized simple, patient-centered terminology when reporting CIMC data. This terminology should be based on evidence and not be overly prescriptive. In addition, researchers and analysts should include previous analysis approaches and terminology when necessary to permit comparisons to data from previous studies of existing methods.</td></tr>
<tr><td>2.2</td><td>When developing instruments to measure CIMCs in trials, it is critical that researchers:<br>a. Follow current recommendations from regulatory agencies and professional societies and consortia, such as the FDA's guidance on PRO[viii] use in trials [1] and the PROTEUS Consortium's recommendations on measuring PROs effectively[ix].<br>b. Use simple, evidence-based patient-centered terminology both within instrument items, prioritizing the perspective of the contraceptive user and simplicity over the convenience of the researcher or imposing definitions or characterizations that may not be sufficiently objective to translate across contexts.[x]<br>c. Ensure instruments are accessible to those with differing literacy levels and people with disabilities, including consideration of pictorial instruments and adherence to current USG plain language guidelines [2] and Web Content Accessibility Guidelines (WCAG) [3] for digital content.<br>Ensure instruments adhere to data privacy requirements, legal protections, and ethical considerations to protect data and prevent use of data by any entity outside of those described in the trial informed consent forms.</td></tr>
<tr><td>4.1</td><td>Presently, researchers should include two types of assessments for primary outcomes of CIMCs:<br>a. Bleeding and spotting days, and</td></tr>
</table>

---

[vii] These standards can include FIGO's System 1 for nomenclature of normal and abnormal uterine bleeding [4], the Global CIMC Task Force's definition of CIMCs [5]. recommendations on bleeding data analyses in contraceptive studies [6], and other terminology harmonization efforts, especially those using methodology to establish consensus and priorities (e.g., Delphi method, Child Health and Nutrition Research Initiative [CHNRI] approach).

[viii] All data on CIMCs in trials are reported by the participant (i.e., it is a patient reported outcome [PRO]) unless the trial is using an indirect quantitative and semi-quantitative approach like an alkaline hematin assay to determine menstrual blood loss from used menstrual products. See 4.6, 5.7b, 6.1, and 6.2 for recommendations on these types of instruments.

[ix] PROTEUS recommendations on measuring PROs effectively with ISOQOL standards [7].

[x] If supported by evidence, an example of this could be asking participants about use/nonuse of menstrual products, instead of asking participants to use and understand the term 'spotting', which may not be relevant in all contexts.

| | |
|---|---|
| | b. Frequency, duration, and volume of bleeding episodes. |
| 4.11 | Presently, trials should measure acceptability and quality of life related to CIMCs to assess the impacts of CIMCs on users and inform contraceptive counseling and user choices. This can be done in the following ways:<br>a. In Phase I, II, and III, researchers should measure participants' self-perceived changes in bleeding and other CIMCs (e.g., whether participants self-define changes as significant). This should be measured over time for Phase II and III.<br>b. In Phase II and III, researchers should measure the physical and psychosocial impacts of CIMCs on quality of life, including burdens and benefits. The aspects that are measured should be based on local priorities and informed by local researchers and advisors, and may include impacts on work, sexual well-being, mental burden, anxiety and stress, quality of relationships, social well-being, and financial well-being (including the burden of menstrual health management).<br>c. In Phase II and III, researchers should measure changes in acceptability related to CIMCs over time. |
| 5.2 | Presently, researchers should use the following definitions during analysis for bleeding episode and for duration, frequency, and volume of bleeding episodes:<br>a. **Bleeding episode**: one or more consecutive days with bleeding and/or spotting, bordered by 2 full days without any bleeding or spotting.<br>b. **Duration**: A bleeding episode can be classified into prolonged or not prolonged.<br>• Prolonged Duration – Bleeding/spotting episode lasting more than 7 days.<br>c. **Volume:** Volume should be classified according to the participant's perception. Volume should be described as "lighter," "usual," or "heavier" as compared to the individual's typical volume when not using contraception.<br>d. **Frequency**:<br>• Absence of bleeding/spotting: no bleeding or spotting during the reference period.<br>• Infrequent bleeding: 2 or fewer bleeding/spotting episodes during a 90-day reference period.<br>• Frequent bleeding: > 4 bleeding/spotting episodes during a 90-day reference period.<br>• Scheduled bleeding: a bleeding/spotting episode starting during the expected interval and lasting no more than 7 days for a method expected to have a predictable bleeding pattern.[xi]<br>• Unscheduled bleeding a bleeding/spotting episode occurring at any time prior to the expected interval, even if it continues into the next expected interval for a method expected to have a predictable bleeding pattern.[xii] |
| 5.3 | Presently, analysts should report the following **variables** on *number of bleeding and spotting days* during the reference period:<br>a. Total number of bleeding or spotting days, |

[xi] Predictable bleeding pattern: an expected predictable pattern is based on the product design as determined by the regimen (e.g., combined hormonal methods [oral, transdermal, vaginal, or injectable] and non-hormonal methods such as IUDs or non-hormonal vaginal products). Unpredictable bleeding pattern: an expected unpredictable pattern based on the product design (e.g., progestin only methods including implants, IUDs, injectables, pills, patches, and rings) [6].

| | |
|---|---|
| | b. Number of bleeding days,<br>c. Number of spotting days,<br>Number of scheduled and unscheduled bleeding or spotting days, if relevant for the method as described in 5.2d. |
| 5.11 | Presently, analysts should report the following about **missing data**[xii] on the number of *bleeding and spotting days*, as well as *frequency, duration and volume of bleeding*, during the reference period:<br>a. For Phase I and IIa trials: The number of discontinuations and the reasons for discontinuation<br>For Phase IIb, III and IV trials: The number of discontinuations and the reasons for discontinuation; and the potential impact of informative drop-out when describing CIMCs over time if a meaningful percentage of trial participants discontinue due to CIMCs[xiii] |

[xii] Data may be missing due to early method discontinuation, trial discontinuation, or loss to follow up.

[xiii] This is to account for the distribution of CIMC outcomes towards the end of the trial being disproportionately represented by subjects who did not experience those outcomes or experienced less impactful outcomes. Approaches could include (a) reporting CIMC outcomes separately among all enrolled participants (where the denominator is the number of subjects still in follow-up at a given time) and among participants who completed the trial (where the denominator is uniform over time), where differences in patterns would suggest that informative drop-out was biasing the results; or (b) explicitly modeling the (potentially informative) drop-out mechanism using an appropriate methodology, such as the one developed and performed in the context of DMPA-induced amenorrhea [8,9] using weighted generalized estimation equations (GEE) for population-level analyses [10].
